# Multi-ancestry GWAS deciphers genetic architecture of abdominal aortic aneurysm and highlights *PCSK9* as a therapeutic target

**DOI:** 10.1101/2022.05.27.22275607

**Authors:** Tanmoy Roychowdhury, Derek Klarin, Michael G Levin, Joshua M Spin, Yae Hyun Rhee, Alicia Deng, Colwyn A Headley, Ida Surakka, Noah L Tsao, Corry Gellatly, Verena Zuber, Fred Shen, Whitney E Hornsby, Ina Holst Laursen, Shefali S Verma, Adam E Locke, Gudmundur Einarsson, Gudmar Thorleifsson, Sarah E Graham, Ozan Dikilitas, Jack W Pattee, Renae L Judy, Ferran Pauls-Verges, Jonas B Nielsen, Brooke N Wolford, Ben M Brumpton, Jaume Dilmé, Olga Peypoch, Laura Calsina Juscafresa, Todd L Edwards, Dadong Li, Karina Banasik, Søren Brunak, Rikke L Jacobsen, Minerva T Garcia-Barrio, Jifeng Zhang, Lars M Rasmussen, Regent Lee, Ashok Handa, Anders Wanhainen, Kevin Mani, Jes S Lindholt, Lasse M Obel, Ewa Strauss, Grzegorz Oszkinis, Christopher P Nelson, Katie Saxby, Joost A van Herwaarden, Sander W van der Laan, Jessica van Setten, Mercedes Camacho, Frank M Davis, Rachael Wasikowski, Lam C Tsoi, Johann E Gudjonsson, Jonathan L Eliason, Dawn M Coleman, Peter K Henke, Santhi K Ganesh, Y Eugene Chen, Weihua Guan, James S Pankow, Nathan Pankratz, Ole B Pedersen, Christian Erikstrup, Weihong Tang, Kristian Hveem, Daniel Gudbjartsson, Solveig Gretarsdottir, Unnur Thorsteinsdottir, Hilma Holm, Kari Stefansson, Manuel A Ferreira, Aris Baras, Iftikhar J Kullo, Marylyn D Ritchie, Alex H Christensen, Kasper K Iversen, Nikolaj Eldrup, Henrik Sillesen, Sisse R Ostrowski, Henning Bundgaard, Henrik Ullum, Stephen Burgess, Dipender Gill, Katherine Gallagher, Maria Sabater-Lleal, DiscovEHR, Regeneron Genetics Center, UK Aneurysm Growth Study, DBDS Genomic Consortium, VA Million Veteran Program, Gregory T Jones, Matthew J Bown, Philip S Tsao, Cristen J Willer, Scott M Damrauer

## Abstract

Abdominal aortic aneurysm (AAA) is a common disease with significant heritability. In this study, we performed a genome-wide association meta-analysis from 14 discovery cohorts and uncovered 144 independent associations, including 97 previously unreported loci. A polygenic risk score derived from meta-analysis was able to explain AAA beyond clinical risk factors. Genes at AAA risk loci indicate involvement of lipid metabolism, vascular development and remodeling, extracellular matrix dysregulation and inflammation as key mechanisms in the pathogenesis of AAA. We further integrated functional data to elucidate expression of genes associated with AAA. These genes also indicate crossover between the development of AAA and other monogenic aortopathies, particularly via TGF-β signaling pathways. Motivated by the strong evidence for the role of lipid levels in AAA by PheWAS, we identified therapeutic opportunities using Mendelian Randomization and, in pre-clinical studies, we demonstrated that *PCSK9* inhibition in mice prevented the development of AAA.

## Introduction

Abdominal aortic aneurysm (AAA) is a life-threatening condition in which progressive expansion of the infrarenal aorta may lead to rupture, which is associated with high mortality. Approximately 4% of the U.S. population over 65 years of age is affected by AAA, resulting in ∼41,000 deaths annually (Stuntz, 2016; Summers et al., 2021).

AAA are often discovered incidentally or as a result of screening programs in certain demographic groups. Current USPSTF guidelines recommend screening via duplex ultrasonography in men aged 65-75 years old who have ever smoked (O’Donnell and Schermerhorn, 2020), since men develop AAA at 3-4 times the rate of women (Lo and Schermerhorn, 2016) and smoking is a key risk factor (Pleumeekers et al., 1995). The mainstay of management is longitudinal surveillance until aneurysm size reaches the point at which the risk of rupture exceeds the risk of repair (Chaikof et al., 2018). This disease surveillance period, which may last several years, represents an ideal opportunity to intervene and prevent disease progression. Unfortunately, there are currently no approved pharmacological therapies for the prevention and treatment of AAA. While multiple pharmacological therapies have been previously proposed, based on compelling biology and promising evidence from preclinical model systems, including ACE inhibitors, angiotensin receptor blockers, matrix metalloproteinase inhibitors, and statins, to date, none have been shown to affect aneurysm growth or rupture in human trials (Chaikof *et al*., 2018).

Over the last 2 decades, large-scale genetic analyses have been instrumental in revealing novel targets and promising therapies for atherosclerotic conditions (Cohen et al., 2006; Dewey et al., 2017). Previous genome-wide association studies (GWAS) of AAA have revealed 24 genomic risk loci for AAA (Bown et al., 2011; Bradley et al., 2013; Gretarsdottir et al., 2010; Jones et al., 2017; Klarin et al., 2020), but a significant portion of AAA heritability remains unexplained. Here, we leveraged genetic data across 17 studies to: 1) perform a genetic discovery analysis for AAA with substantially higher case numbers than previous studies (5-fold increase); 2) create and test the predictive power of polygenic risk score derived from this analysis; 3) prioritize causal genes and pathways leading to disease; 4) explore the spectrum of phenotypic consequences associated with AAA risk variants; and 5) identify potential therapeutic targets which may help prevent and treat AAA.

## Results

### Multi-ancestry meta-analysis identifies 97 novel risk loci

To identify genetic variants associated with AAA, we performed a meta-analysis of 17 individual GWAS from 14 discovery cohorts in the AAAgen consortium (**Figure 1a, Table S1, Figure S1-2**). Our analysis was comprised of 39,221 individuals with AAA (37,214 of European (EUR) ancestry and 2,007 of African (AFR) ancestry). After meta-analysis we obtained single variant association statistics for 55.8M variants, of which 33.4M were present in two or more GWAS and were used for downstream analyses. We identified 126 index variants associated with AAA at a genome-wide significance threshold (P<5×10^−8^) (**Figure S3, Table S2**). None of the index variants displayed significant evidence for heterogeneity (Heterogeneity test; P>0.05/126) of effect estimates among the contributing GWAS (**Table S2**). We observed consistent effect size estimation for index variants in a comparison between meta-analysis with or without summary statistics of AFR ancestry (**Figure S4**). Since approximately 45% of all cases were contributed by the VA Million Veteran Program (MVP) EUR analysis, we performed another meta-analysis without this cohort and tested for nominal significance (P<0.05) in both datasets as internal replication. Among 126 index variants, 3 rare variants (minor allele frequency (MAF) <0.01) failed to meet our internal replication threshold. (**Figure S5, Table S2**). These 3 loci, along with 2 other loci with rare (MAF<0.01) index variants, were not further investigated. Of the remaining 121 genome-wide significant loci, 97 were not previously reported (**Figure 1b**). We replicated all 24 loci that were reported previously as associated with AAA (Klarin *et al*., 2020) with P< 5×10^−8^. The index variants represented a wide spectrum of allele frequencies, with 6 being low allele frequency (MAF 0.01-0.05) and the rest common (MAF > 0.05). As expected, by substantially (∼5-fold) increasing the number of participants with AAA compared to previous reports (Klarin *et al*., 2020), we were able to identify variants with lower effect estimates (**Figure 1b**) that could not be discovered with smaller sample sizes. Using approximate conditional analysis, we identified 23 additional genome-wide significant variants (**Table S3**) within the associated regions that were statistically independent of the 121 index variants. This resulted in 144 statistically independent associated variants.

**Figure 1:**
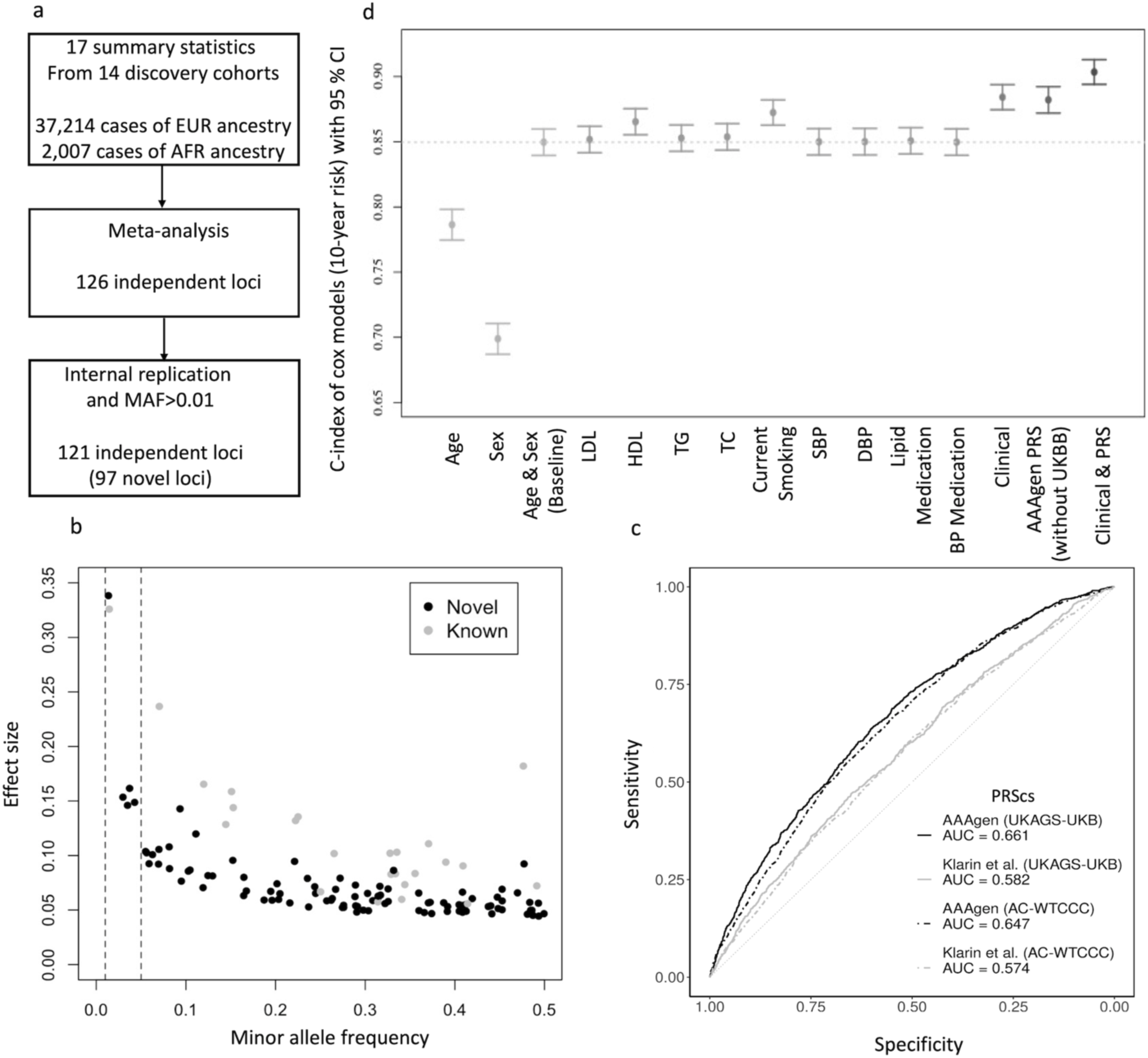
GWAS meta-analysis and PRS of AAA. **a)** Flowchart of GWAS meta-analysis. The initial analysis generated 126 genome-wide significant loci but 5 were excluded based on internal replication strategy and minor allele frequency (MAF) threshold. **b)** Minor allele frequency is plotted against effect estimates for genome-wide significant index variants. The robust increase in sample size compared to previous studies allowed for the identification of novel disease associated variants with smaller effect estimates. Two dotted lines represent MAF 0.01 and 0.05. **c)** Performance of PRS constructed based on the current meta-analysis (AAAgen) was compared with Klarin et al. (Klarin *et al*., 2020), the largest previously published GWAS of AAA. We observed improved prediction by area under the curve (AUC) in both validation datasets. **d)** C-index of cox models (10-year risk) with 95% confidence intervals (CI) in UKBB. The baseline model includes age, age^2^ and sex. The dashed line is at C-index value from the baseline model. All subsequent models with clinical measurements and PRS incorporate the baseline variables.

### Polygenic risk scores explain AAA beyond clinical risk factors

To evaluate the ability of our meta-analysis to explain observed disease and potentially predict future disease we generated weights for polygenic risk scores (PRS) using PRScs (Ge et al., 2019), which calculates posterior SNP effect estimates from original GWAS effect estimates using a Bayesian approach. For comparison, we generated weights from Klarin et al. (Klarin *et al*., 2020) (7,642 cases), the largest GWAS of AAA prior to this study, using the same methodology. These two sets of weights were then used to calculate PRS in two external validation cohorts that had not been included in the discovery study to estimate improvement in AAA risk assessment with the five-fold increase in case number in this meta-analysis. Not surprisingly, the AAAgen PRS (AUC; UKAGS-UKB: 0.66, AC-WTCCC: 0.64) outperformed that of Klarin et al. (Klarin *et al*., 2020) PRS (AUC; UKAGS-UKB: 0.58, AC-WTCCC: 0.57) in both the validation cohorts (**Figure 1c, Figure S6**), explaining an additional 13% to 14% of disease associated variance in these cohorts.

To further evaluate the predictive power of the 2022 PRS on incident AAA we conducted analyses in data from UK Biobank (UKBB). To avoid overfitting, we performed another meta-analysis without UKBB summary statistics and generated weights for PRS as described above. We used UKBB hospital and cause of death registry data in longitudinal Cox Proportional Hazards models. First, we tested the predictive performance of a baseline model including age, age^2 and sex to establish a point of comparison for the predictive power of different clinical risk factors and the AAAgen PRS (without UKBB). When comparing Harrel’s C-index (**Figure 1d, Table S4**) from the model where PRS was added to the baseline predictors (C-index = 0.850, 95% Confidence Interval [0.840; 0.860]), the predictive power of the PRS (C-index = 0.882 [0.872; 0.892]) was better than that of the most predictive clinical risk factors, including smoking status (C-index = 0.872 [0.863; 0.882]), and similar to the model including all tested clinical factors together (C-index = 0.884 [0.875; 0.894]). Furthermore, adding both the PRS and all clinical factors into one model yielded a C-index of 0.904 [0.894; 0.913], which represents remarkably high concordance between predicted and observed cases in a population-based cohort with notable selection bias towards healthy individuals, and substantial improvement over baseline demographics only (Δ = 0.054).

### Enrichment analyses highlight relevant biological functions, tissues and cell types

We performed a gene-set enrichment analysis using reconstituted gene-sets in DEPICT (Pers et al., 2015). Out of 14,462 reconstituted gene-sets, 852 were significant at FDR < 0.05 (**Table S5**). Among the top 30 gene-sets, we primarily observed enrichment of several terms associated with genesis, morphogenesis, development, proliferation, and migration of elements in the cardiovascular system (**Figure 2a**). Notably, collagen and TGF-β binding were highlighted as enriched gene-sets.

**Figure 2:**
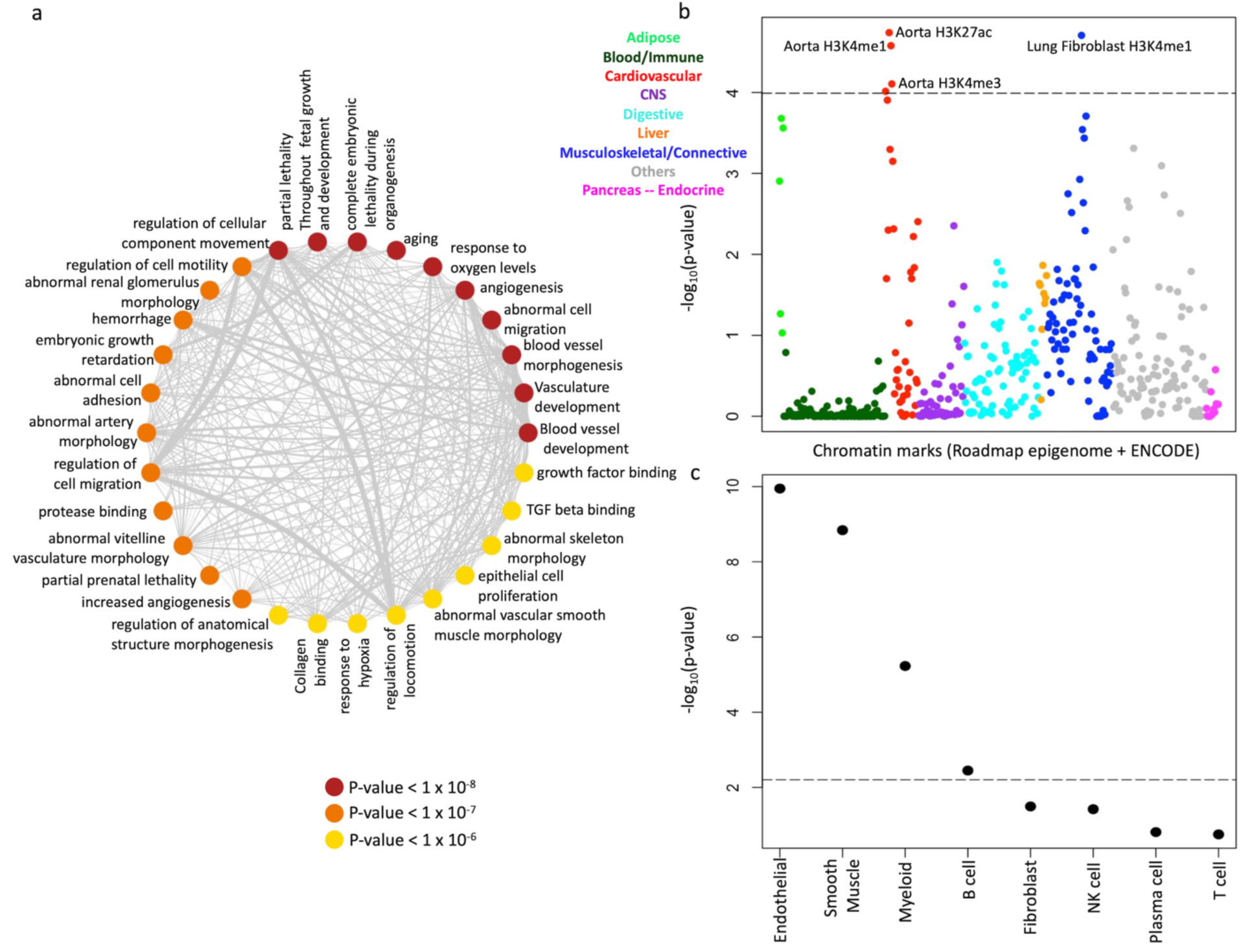
Enrichment analysis. **a)** Gene-set enrichment analysis by DEPICT. Nodes represent the top 30 gene-sets prioritized by DEPICT (colored by p-value). Thickness of edges represent overlap between gene-sets. **b)** P-values for enrichment of per-SNP heritability calculated by LDSC using tissue specific chromatin marks. Different colors were used to classify tissues from broad categories. The dotted line represents the significance threshold after correcting for multiple testing. **c)** P-values for estimation of non-zero regression coefficient for each cell-type calculated by RolyPoly using scRNA of aorta. The dotted line represents the significance threshold after correcting for multiple testing.

To understand the causal tissues where genes in the GWAS loci might be functional, we employed a stratified LD score regression (LDSC) (Finucane et al., 2018) analysis using histone marks from ENCODE and Roadmap Epigenomics. This analysis identified enrichment of per-SNP heritability in H3K27ac (P=1.8×10^−5^), H3K4me1 (P=2.6×10^−5^) and H3K4me3 (P=7.8×10^−5^) marks in aorta (**Figure 2b**). This indicates significant involvement of aortic tissue biology downstream of the observed GWAS signals.

Variants identified by GWAS often affect genes that are only active in a subset of cell types in a tissue. To further investigate the involvement of particular cell types of aorta, single cell RNA (scRNA) (**Figure S7**) from aorta (Davis et al., 2021) was analyzed using RolyPoly (Calderon et al., 2017). This regression-based polygenic model uses GWAS summary statistics and scRNA data to learn a regression coefficient that captures each cell type’s influence on the variance of GWAS effect estimates. Using regression coefficients, 4 out of 8 cell types were found to be enriched (P<0.05/8), with endothelial (P=1.1×10^−10^) and smooth muscle (P=1.4×10^−9^) cells being most strongly associated with AAA (**Figure 2c**). The involvement of endothelial cells likely highlights the overlap with atherosclerosis whereas the smooth muscle involvement is consistent with the medial degeneration typical of AAA pathogenesis.

### Gene-prioritization identifies putative causal genes at AAA risk loci

We used 8 different indicators to prioritize a single putative causal gene at the 121 genome-wide significant loci. For this, candidate genes were selected using the following indicators: 1) A gene with protein-altering variant that is either the index variant or in high LD (>0.8) with the index variant. We observed 21 loci with such genes (**Table S2**). 2) An eQTL colocalization or significant association in transcriptome wide association study (TWAS) using GTEx v8 (Consortium, 2020) eQTL data from 4 tissues: aorta, liver, whole blood and adipose. These analyses identified 82 and 75 loci with at least one gene prioritized by colocalization and TWAS respectively (**Table S6-7**). 3) A gene prioritized by Polygenic Priority Score (PoPS) (Weeks et al., 2020), a similarity-based method that leverages publicly available functional genomics resources. PoPS was developed with the assumption that causal genes share similar functional characteristics. For each locus, we ranked the genes within 1 Mb (either direction) of the index variant and reported the gene with the highest score as the gene prioritized by PoPS. Out of 121 loci, 111 loci had at least one gene with a score in the top 10% of PoPS (**Table S8**). 4) A gene with evidence of causing a relevant monogenic phenotype using ClinVar and Renard et al. (Renard et al., 2018) within 1 Mb of the index variant (**Table S9-10**). We found 48 loci with at least one such gene. 5) A gene that is closest to the index variant (**Table S2**).

In total, we obtained 523 candidate genes from 121 loci using 5 above indicators. Next, these genes were queried in 3 other datasets to obtain additional prioritization evidence. 1) We utilized a trio of datasets [(Maegdefessel et al., 2014; Spin et al., 2011) and GSE197748] examining gene transcriptional profiling in two AAA models (i.e. ApoE-/-/AngII and porcine pancreatic elastase-PPE), specifically those comparing sham/control aortic gene expression vs. expression in AAA aortic tissue in experiments featuring 10-week-old male and female mice. Of the 523 candidate genes, 214 demonstrated differential expression in the mouse AAA gene set (**Table S11**). 2) Expression in human abdominal aortic tissue (AAA vs control) from Biros et al. (Biros et al., 2015) where 79 genes with q-value<0.05 were included as supported by this indicator (**Table S12**). 3) Related phenotypes observed in mouse knock-outs for candidate genes. We queried data from the Mouse Genome Informatics database and International Mouse Phenotyping Consortium to identify genes for which knockout mice had been reported to have aortic phenotypes consistent with AAA. This generated additional support for 5 genes in this approach (**Table S13**).

Next, we used the following rules, in order of precedence, to prioritize single likely causal genes in 121 loci. 1) At 21 loci, genes with protein-altering variants were prioritized. 2) At 52 loci, we could assign a single likely causal gene by consensus i.e, a single gene was supported by more indicators (minimum 3) than other genes at that locus. At 11 loci, we observed more than 1 gene with same number of indicators (≥3). We used distance from index variant as a tie-breaker to prioritize single genes at these loci (e.g. *SMAD3* was prioritized over *SMAD6*). The above-mentioned steps prioritized a single gene at 84 loci (**Figure 3a, Table S14**). 3) In the remaining 37 loci with no clear consensus candidate gene, we prioritized the closest gene from the index variant as the likely causal gene, as this has been shown to perform reasonably well (∼65-70%) in recent studies (Mountjoy et al., 2021; Pietzner et al., 2021).

**Figure 3:**
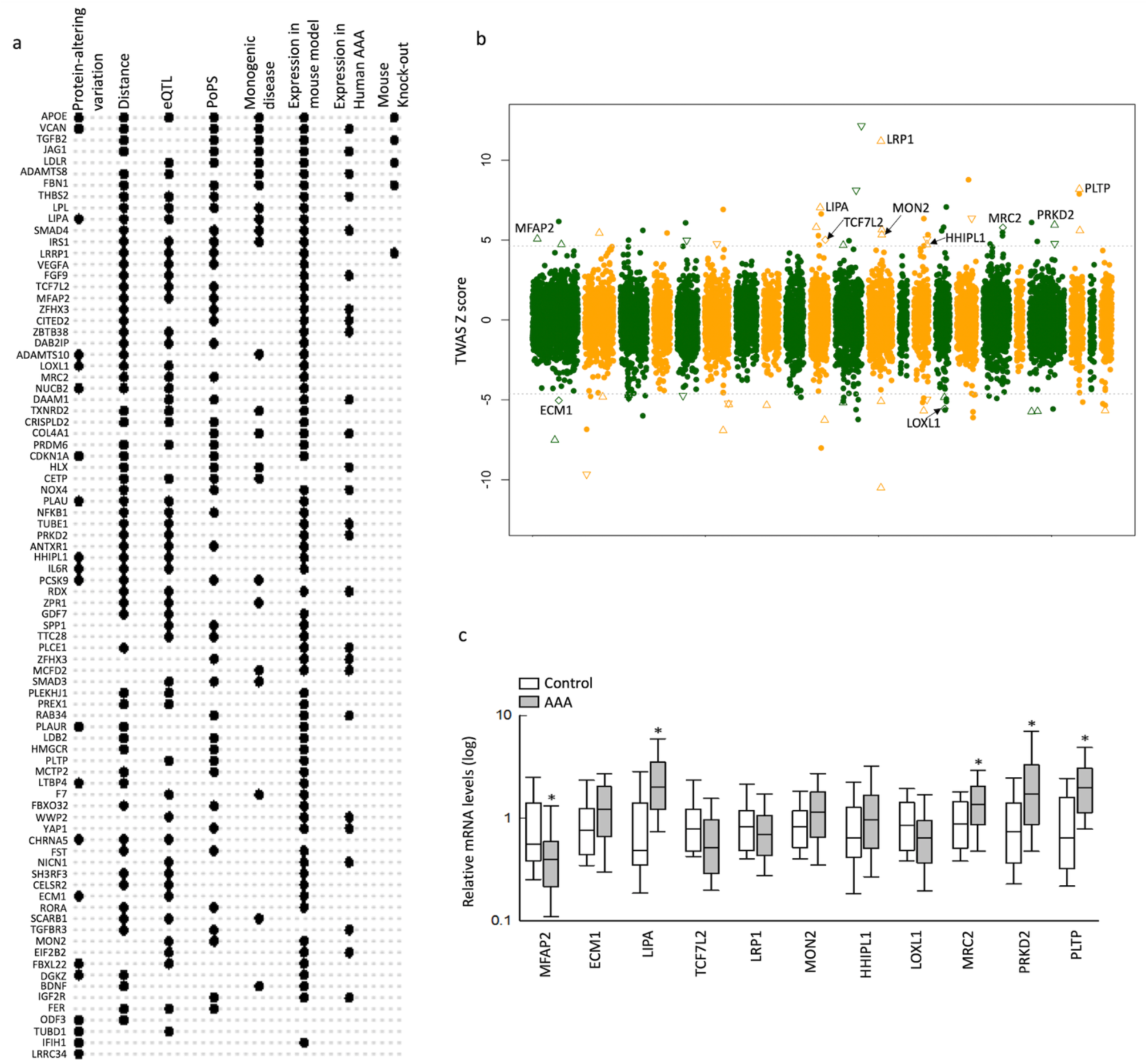
Gene prioritization and expression. **a)** Support of various gene prioritization indicators for 84 loci where a putative causal gene could be prioritized by protein-altering variants or by consensus. Rows (gene names) represent these loci and columns represent 8 supporting indicators used for the prioritization. Black dots at row/column intersection indicate support by a particular indicator for a particular gene. **b)** Aortic tissue-based TWAS z-scores plotted against genomic coordinates of genes. The dotted line represents the associated z-score for significance threshold after multiple testing correction. A positive or negative z-score indicates the association of respectively higher or lower gene expression with AAA. Triangle/square(s) represent genes that were also differentially expressed in the mouse model of AAA. Up/downward triangles represent genes that were observed to have high and low expression respectively in the mouse model. Squares represent genes where both directions were observed. Eleven prioritized genes (see Results) are highlighted with text. **c)** Results of qPCR in 11 prioritized genes. * indicates 5 genes with a significant (after multiple testing correction) difference in expression level between cases and controls.

Next, these 121 prioritized genes, and an additional 12 genes **(Table S3)** that were the gene closest to secondary signals at GWAS loci, were used to identify enriched gene ontology (GO) terms (**Table S15**) using Enrichr (Kuleshov et al., 2016). In GO molecular functions, consistent with previous knowledge regarding the importance of atherosclerosis in AAA, we observed enrichment of genes related to lipid metabolism, including genes involved in binding lipoprotein particle/receptor(s) (*APOE, LDLR, LPL, PCSK9, PLTP, SCARB1*). We also observed enrichment of TGF-β related genes (*TGFBR3, TGFB2, SMAD3, TGFB3, TGFBRAP1, LTBP4, GDF7*) that are likely involved in vascular development. TGF-β related genes, as well as several other prioritized genes (*IL6R, VEGFA, INHBA, IL1F10*), are also involved in cytokine activity and receptor binding, as key modulators of inflammatory mechanisms. We observed 7 genes that encode transcription factors (*HDAC7, LDB2, RBBP8, SMAD3, TCF7L2, WWP2, YAP1*) and 3 additional transcriptional co-activators (*RORA, SMAD4, TERT*). A study of murine and human cardiac fibrosis determined that *WWP2* was a master regulator expressed in diseased human heart to turn on pro-fibrotic and extracellular matrix genes, and was induced by TGF-β and SMAD related pathways (Chen et al., 2019). Other studies showed that *YAP1* is a central regulator of phenotypic switching of vascular smooth muscle cells (VSMCs) (Xie et al., 2012) and negatively regulates differentiation of VSMCs from cardiovascular progenitor cells by decreasing transcription of myocardin (Wang et al., 2017b). Eight prioritized genes were highlighted for growth factor activity/receptor binding (*JAG1, VEGFA, FGF9, BDNF, INHBA, PDGFRA, FER, IL6R)* which are known to be involved in vascular remodeling. The most significantly enriched GO cellular component term was the collagen-containing extracellular matrix (*SEMA7A, TGFB2, ECM1, TGFB3, LTBP4, THBS2, LOXL1, ADAMTS10, F7, VCAN, COL4A1, MFAP2, COL21A1, APOE, ADAMTS8, FBN1)* indicating significant involvement of extracellular matrix dysregulation in AAA pathogenesis.

### Prioritized genes indicate genetic overlap with thoracic aortic aneurysm

While there is human genetics evidence regarding the role of TGF-β signaling pathway in thoracic aortic aneurysm (TAA) (Pinard et al., 2019), which is characterized by dilation of the aortic root or the ascending/descending aorta nearest the heart itself, this GWAS provides the human genetics evidence of the involvement of this pathway in AAA as well, and suggests shared biology between the two diseases. To further investigate the overlap between AAA and TAA genes, we queried the association of AAA index variants in a recent TAA GWAS (n=1,351) (Roychowdhury et al., 2021) and observed 24 AAA index variants associated with TAA at P<0.05. These variants are generally associated with AAA with weaker effect estimates compared to TAA (**Figure S8**). This observation is consistent in two genome-wide significant common variant associations of TAA intronic to the *FBN1* (coloc; PP4: 0.94) and *TCF7L2* (coloc; PP4: 0.92) (**Figure S8**) as well. While 7 of 24 abovementioned variants (at *ECM1, DNM3, TCF7L2, CMIP, RAB34, GDF7, VCAN*) are also associated with lipid traits, the effect estimates for lipid traits are relatively weaker than either type of aneurysm for these variants (**Table S16**).

Unlike AAA, ∼10-20% of TAA cases have a monogenic origin (Beil et al., 2021). Supported by varying levels of evidence, Renard et al. (Renard *et al*., 2018) compiled a list of 53 genes that may cause monogenic TAA because of single pathogenic variants. Many of these genes represent pathways (e.g., TGF-β signaling) involved in vascular development and/or extracellular matrix organization. Among Renard’s 53 genes, we observed a genome-wide significant variant for AAA within 1 Mb of 14 (*FBN1, ACTA2, SMAD3, SMAD4, SLC2A10, PKD2, TGFB2, HCN4, JAG1, ADAMTS10, TGFB3, COL4A1, VCAN, SMAD6*). Of note, 12 of the 14 genes were supported by at least 3 indicators in the gene prioritization step and 9 (*FBN1, SMAD3, SMAD4, TGFB2, JAG1, ADAMTS10, TGFB3, COL4A1, VCAN*) were prioritized as the likely causal genes. While the contribution of the above-mentioned pathways is evident in both AAA and TAA, stronger effect estimates or monogenic mechanisms likely indicate a stronger contribution to TAA compared to AAA. In contrast, differences in overlap with lipid metabolism genes likely indicate larger involvement of lipid levels as a risk factor for AAA compared to TAA. Consistent with this, previous studies reported a significant genetic correlation between lipids and AAA (Klarin *et al*., 2020), but not TAA (Roychowdhury *et al*., 2021).

### Integration with external datasets elucidates aortic expression of AAA associated genes

Most (87%) of the prioritized genes are expressed in aorta, as observed in RNA-seq of abdominal aorta from AAA patients (**Figure S9**). In scRNA data of aortic tissue, as predicted by RolyPoly analysis, the prioritized genes are primarily expressed in endothelial and smooth muscle cells (**Figure S9**).

GWAS often identify non-coding variants that are hypothesized to be associated with disease via alteration of gene expression levels of causal genes. To further dissect this possibility, TWAS methods leverage a predictive model of gene expression from a reference panel (eQTLs) to predict gene expression from GWAS summary statistics, followed by a test of association between predicted expression and phenotype (Wainberg et al., 2019). Accordingly, we performed a TWAS using aortic tissue reference panel (Consortium, 2020) and identified 104 genes for which predicted gene expression was associated with AAA (**Table S6**). To validate the TWAS analyses, we further inquired whether genes identified by TWAS were differentially expressed in aortic tissue from mouse models of AAA (**Table S11**). Of 45 genes that were present in both sets, 19 genes were differentially expressed in the same direction during mouse AAA development and TWAS prediction. Additionally, 4 genes displayed mixed direction in the mouse model, i.e., direction matched with TWAS for some experimental conditions. Out of the 23 genes identified in TWAS that showed differential expression in mouse AAA models, 11 (**Figure 3b**) were prioritized as likely causal genes by consensus.

We further validated the 11 genes identified on TWAS with supportive mouse gene expression data in human AAA tissue using qPCR (Sola-Villa et al., 2015) in 97 individuals with AAA and 36 without AAA. Five of these 11 genes (**Figure 3c, Table S17**) were differentially expressed (P<0.05/11). Whereas all 5 genes were up-regulated in both the TWAS prediction and in the mouse data, 4 genes (*LIPA, MRC2, PRKD2* and *PLTP*) were up-regulated while 1 (*MFAP2*) was down-regulated, when comparing expression in aneurysmal to non-aneurysmal abdominal aortic tissue. Two of these five genes (*LIPA* and *PLTP*) likely act through lipids and atherosclerosis. Of the remaining genes, *MRC2* functions via extracellular matrix remodeling in conjunction with urokinase plasminogen activator and its receptor (Behrendt et al., 2000) (*PLAU* and *PLAUR*, both of which were also prioritized by the GWAS); a microfibrillar glycoprotein (*MFAP2*) known to bind to *FBN1* and *FBN2* (Hanssen et al., 2004); and a serine/threonine protein kinase (*PRKD2*) involved in regulation of cell proliferation via inhibition of HDAC7 (Wang et al., 2022), a gene also prioritized by the GWAS. Overall, our strategy of using genetically controlled expression levels as priors helped us to validate gene expression changes that are likely causally associated with AAA from expression changes due to disease status.

### Pleiotropic AAA risk variant associations identify a significant shared heritability to lipoprotein biology

To assess the pleiotropy associated with AAA risk variants and identify conditions with shared genetic risk, we performed PheWAS of the 121 genome-wide significant index variants against the MRC-IEU open GWAS project (Elsworth et al., 2020). Based on follow-up network analysis (see Methods) using PheWAS summary statistics, we identified 7 distinct modules of phenotype-clusters (**Figure 4a, Table S18**) representing LDL cholesterol, apolipoprotein B, coronary heart disease, anthropometric traits, apolipoprotein A/metabolic biomarkers, blood cell traits and total cholesterol/cardiovascular disease medications. Next, we specifically tested the association of AAA index variants with established clinical risk factors for AAA (Klarin et al., 2018; Klarin *et al*., 2020) using the largest available GWAS summary statistics of 4 lipids traits (Graham et al., 2021), 3 blood pressure traits (Evangelou et al., 2018) and 2 smoking traits (Liu et al., 2019). We observed 52 variants (42 lipids, 11 blood pressure, and 2 smoking) that were genome-wide significant (P<5×10^−8^) in at least one of these traits (**Figure 4b, Table S16**). To elucidate if the same variants are associated with AAA and the risk factors, pairwise colocalization between AAA and each risk factor traits were performed. At a threshold of PP4 > 0.5, we observed colocalization of 33 AAA loci with lipids (including *PCSK9*, PP4=1 for both LDL and total cholesterol), 11 loci with blood pressure and 1 with smoking traits (**Table S19**). This set of analysis highlights that a substantial proportion of AAA loci likely function through modulating blood lipid levels, which in turn contribute to AAA development.

**Figure 4:**
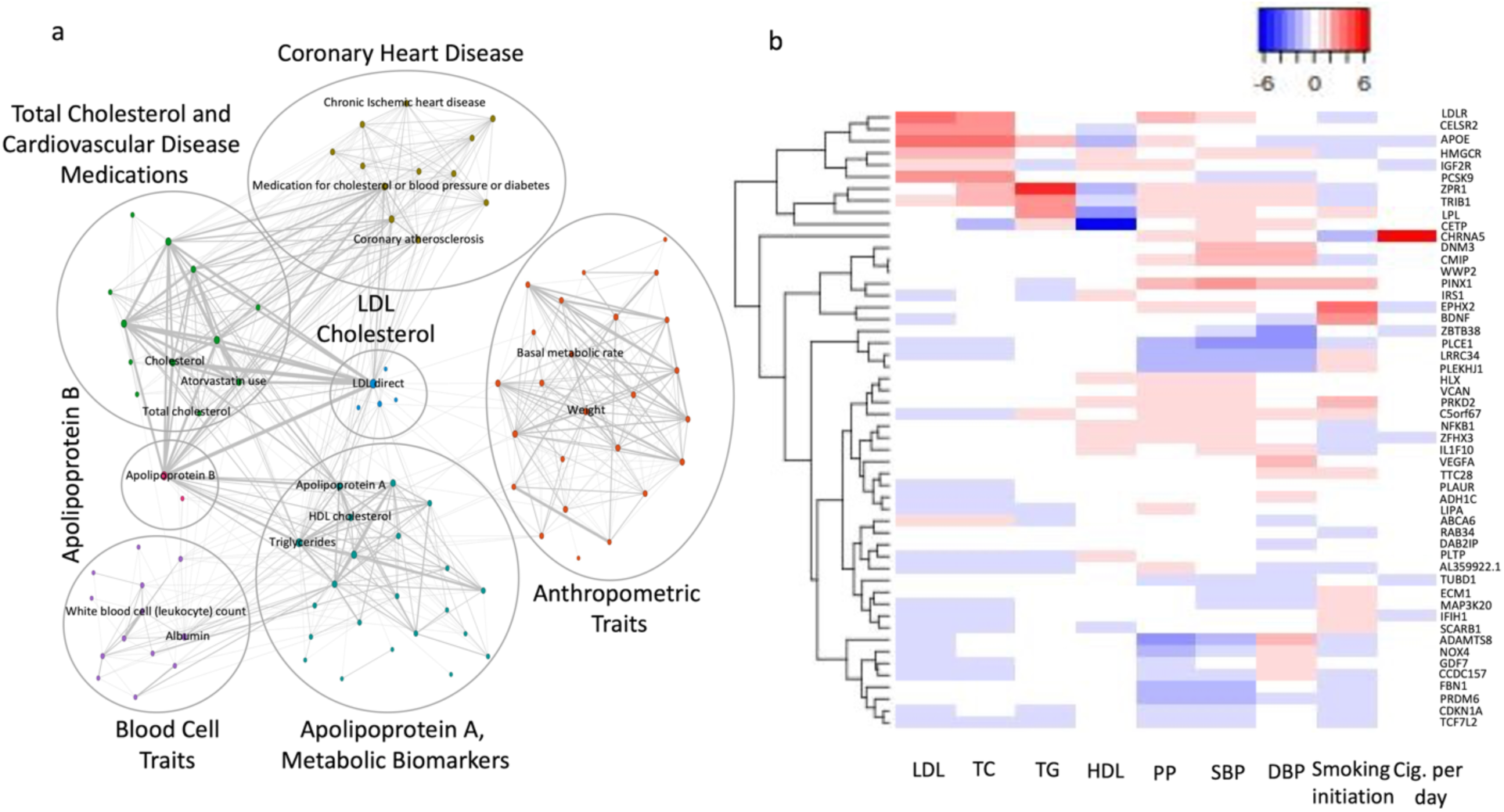
Pleiotropy. **a)** PheWAS network diagram representing 7 modules (indicated by circles). **b)** Hierarchical clustering of scaled, normalized effects (beta/standard error) in established risk factor traits for 52 index variants (see Results). Alleles in risk factor traits were aligned with the AAA increasing allele. Index variants (rows) are named by the prioritized genes.

### Genetic causal inference identifies therapeutic opportunities for lipid modulating therapies

Given the strong contribution of lipid biology to the pathogenesis of AAA identified in our gene prioritization and PheWAS analyses, we next sought to prioritize the role of circulating lipoproteins on AAA risk. We first performed conventional inverse variance weighted Mendelian randomization, finding associations between each major lipoprotein-related trait (nonHDL-C, LDL-C, HDL-C, triglycerides, ApoA1, and ApoB) and AAA (**Figure S10, Table S20-21, Supplementary Methods**). Given the substantial overlap in risk variants associated with each lipoprotein trait, we next performed a Mendelian Randomization Bayesian Model Averaging (MR-BMA) analysis (Zuber et al., 2020), a recently developed analytic tool that applies Bayesian principles to prioritize causal risk factors among correlated exposures (in this case, lipoproteins). MR-BMA generates a marginal inclusion probability that prioritizes causal risk factors for disease, rather than determining effect estimates for each of the lipoproteins on AAA risk, which we have performed previously (Harrison et al., 2018; Klarin *et al*., 2018). Genetic instruments were constructed from independent genetic variants associated with any major lipoprotein-related trait (nonHDL-C, LDL-C, HDL-C, ApoA1, ApoB, or triglycerides) at a genome-wide significance level (P<5×10^−8^, r^2^ < 0.001) in the UK Biobank based on 361,194 EUR ancestry participants, as previously described (Levin et al., 2021) (**Table S22**). Of note, given the substantial correlation between the nonHDL-C and LDL-C lipid fractions (r^2^ > 0.9), two separate models were used for analysis: one containing LDL-C, HDL-C, ApoA1, ApoB, and triglycerides (n = 519 independent genetic variants), and another substituting non-HDL-C for LDL-C (n = 450 independent genetic variants) (**Figure 5a**).

**Figure 5.**
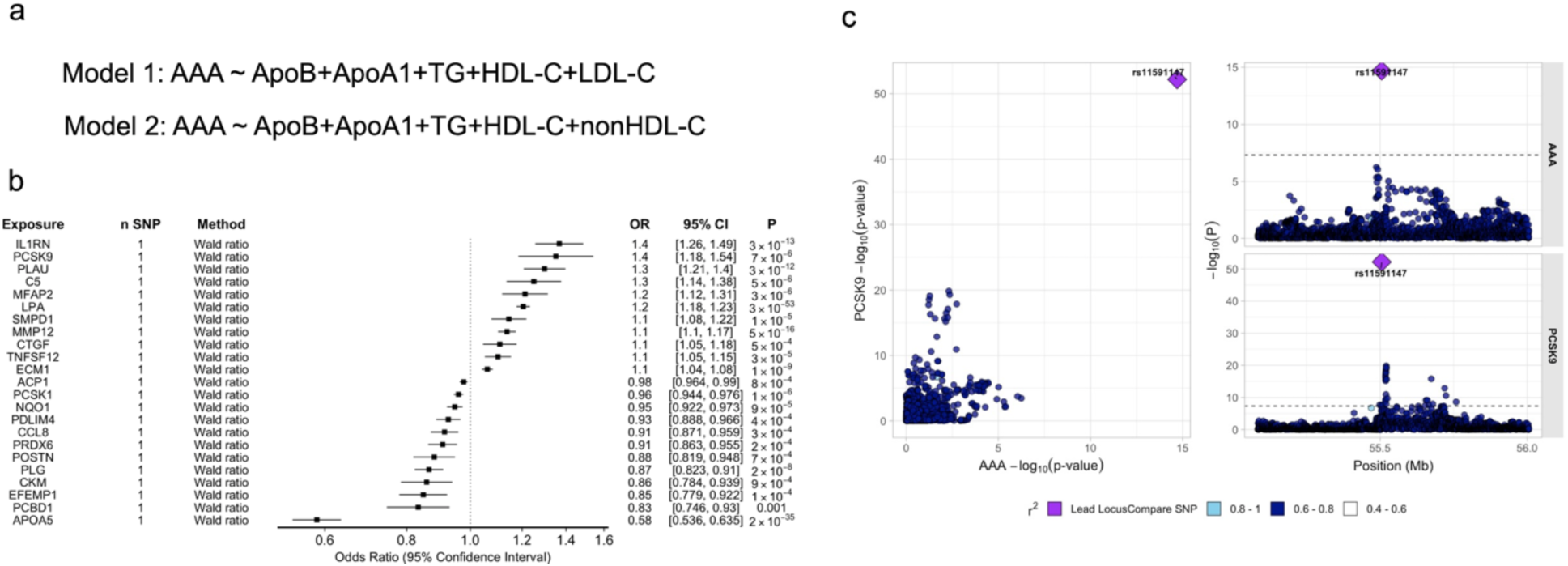
Causal inference methods for the evaluation of AAA risk. **a)** In the MR-BMA analysis to prioritize likely causal lipoprotein fractions, two separate models were used for analysis given the strong correlation between nonHDL-C and LDL-C. All other lipid fractions were included in both models. **b)** A proteome-wide Mendelian randomization analysis using high confidence cis-acting genomic instruments for circulating plasma proteins identified 23 putative causal protein-AAA associations at Benjamini-Hochberg FDR P < 0.05. Odds ratios are depicted per unit change in protein level (1 standard deviation). **c)** LocusCompare plot of circulating PCSK9 protein pQTL results and AAA GWAS results demonstrating evidence of colocalization. Abbreviations: LDL-C, LDL Cholesterol; HDL-C, HDL Cholesterol; TG, Triglyceride-rich lipoprotein

LDL-C and nonHDL-C emerged as the most highly prioritized causal lipoproteins for AAA risk (LDL-C marginal inclusion probability = 0.97, P = 0.0004; nonHDL-C marginal inclusion probability = 0.99, P = 6×10^−5^, **Table S23a)**. When comparing models containing LDL-C vs. nonHDL-C variants, r^2^ and BIC values demonstrate better model fit and variance explained for nonHDL-C. This finding suggests that the additional remnant cholesterol contained within the nonHDL-C subfraction is contributing to AAA risk beyond the LDL-C component (**Table S23b)**. This observation is consistent with prior evidence in atherosclerotic coronary artery disease (Helgadottir et al., 2016). There was reduced evidence that ApoA1 containing particles (ApoA1 or HDL-C) also potentially contributed to AAA (marginal inclusion probability = 0.66/0.85, P = 0.01/0.004).

We next sought to leverage our genetic data to screen for possible novel therapeutic targets for AAA. We used a recently developed proteome-wide MR technique using high confidence (“Tier-1” as defined by Zheng et al (Zheng et al., 2020)) cis-acting genomic instruments for circulating plasma proteins that passed consistency and pleiotropy filters. After restricting to 717 circulating proteins with available proteomic and corresponding genomic data, we performed a protein quantitative trait locus (pQTL) screen and set a Benjamini-Hochberg FDR P < 0.05 for statistical significance. In total, we identified 23 putatively causal protein-AAA associations, all of which also demonstrated evidence of colocalization as required by Zheng et al. (Zheng *et al*., 2020) (**Figure 5b**,**c Table S24)**. Notably, we observed that higher genetically predicted circulating PCSK9 (Proprotein convertase, subtilisin/kexin-type 9) and Lp(a) (Lipoprotein(a)) were associated with increased AAA risk; this was supported by evidence of significant colocalization between PCSK9 pQTL and AAA GWAS at the *PCSK9* locus (PP=1). In addition, higher genetically predicted circulating *APOA5* (Apolipoprotein A5) was associated with decreased AAA risk. Hyperlipidemic mice overexpressing *Apoa5* have been shown to have markedly decreased circulating remnant lipoprotein particles (Grosskopf et al., 2012; Mansouri et al., 2008) and individuals with rare protein-altering variants in *APOA5* were observed to have increased levels of remnant cholesterol (Jorgensen et al., 2013), providing further evidence of the effects of remnant cholesterol on AAA risk.

Of the available lipid modulating therapies, statins are currently recommended for patients with AAA to prevent major adverse cardiac events (Chaikof *et al*., 2018), and ezetimibe remains largely understudied in the setting of AAA. Whether these therapies influence AAA development/progression remains uncertain. Triglyceride-rich lipoprotein/ApoA1 modulating agents targeting the lipoprotein lipase or cholesterol ester transfer protein pathways remain under development. Accordingly, we focused on LDL cholesterol reduction strategies through *PCSK9* inhibition for further investigation. Of note, we investigated whether a reduction in AAA risk through genetic *PCSK9* inhibition was maintained regardless of initial LDL cholesterol, in an effort to define a possible “floor” for lipid lowering therapy. We observed a linear relationship between LDL-C reduction through *PCSK9* and AAA risk (Nonlinearity P > 0.2, **Figure S11**), and further details of this analysis are described in the supplementary materials.

### *PCSK9* inhibition and AAA risk in a murine model

*PCSK9* inhibition presents an attractive therapeutic for AAA given the potency of its LDL cholesterol reduction and favorable side-effect profile (Sabatine et al., 2017). Using a porcine pancreatic elastase (PPE) infusion model of AAA in C57BL/6J mice (**Figure 6**), we investigated whether *Pcsk9* null mice demonstrated attenuated aneurysm growth compared to their wild type counterparts. 10-week-old mice received aortic PPE infusion as previously described (Maegdefessel et al., 2012). We observed a significant decrease in expansion of the abdominal aortic diameter (AAD) from day 10 until day 28 for *Pcsk9* null mice compared with wild type mice with brightness modulation (B-mode) ultrasound imaging performed 3, 7, 14, 21, and 28 days after PPE infusion (**Figure 6**, Welch’s t-test *P<0.05 on day 10; **P<0.01 on day 28 for *Pcsk9*^-/-^ vs wild type). As with previous studies (Ioannou et al., 2022; Mbikay et al., 2015; Wang et al., 2017a) *Pcsk9*^-/-^ mice had lower plasma cholesterol, LDL and HDL levels compared to wild type C57BL/6J mice (**Table S25**).

**Figure 6.**
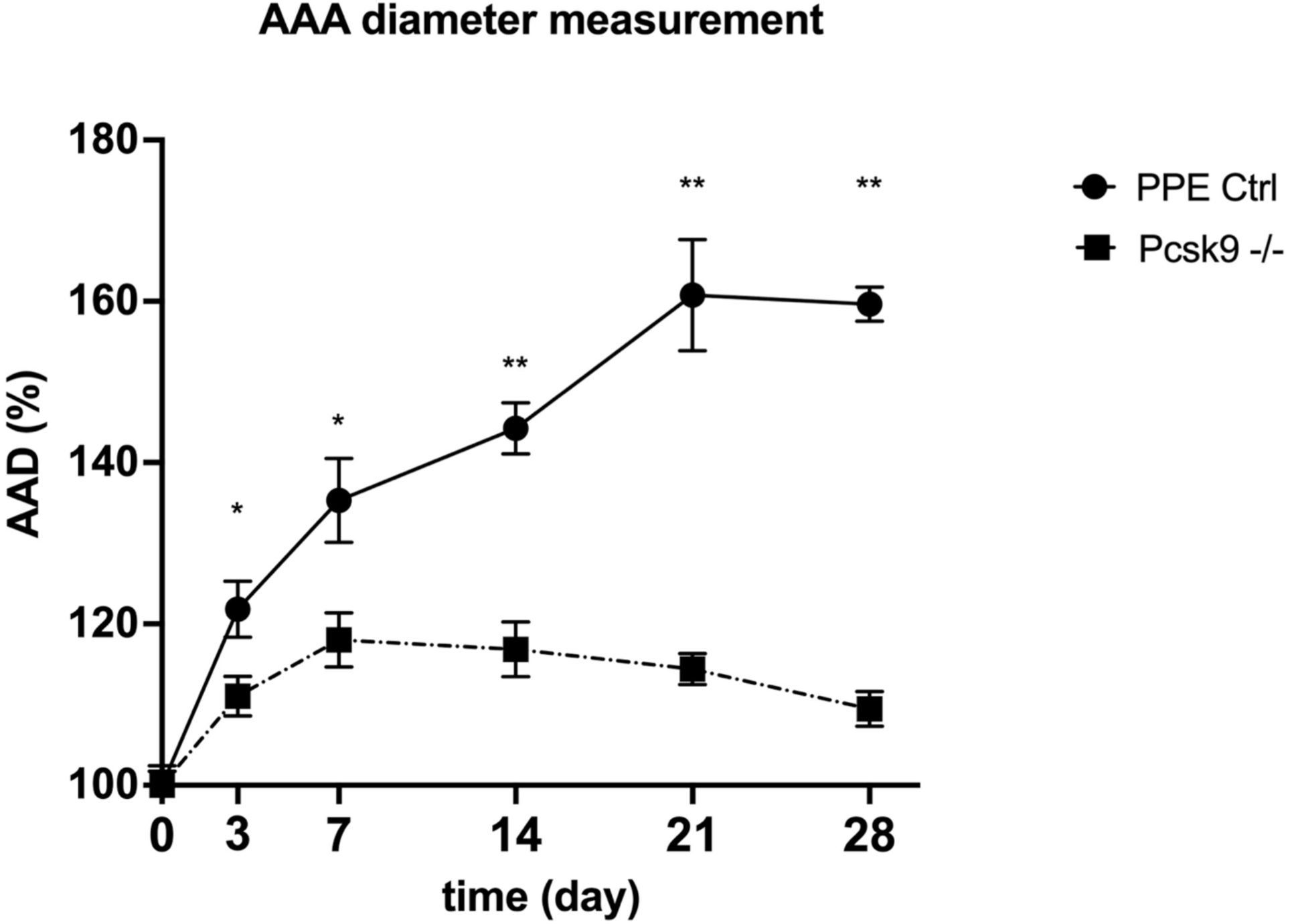
Loss of Pcsk9 inhibits growth of experimental AAA in murine models. PPE surgery was performed on day 0 and maximal aortic diameter was monitored by B-mode ultrasound. Knockout of *Pcsk9* resulted in blunted AAA growth starting at experimental day 3 (PPE Ctrl group n=8; Pcsk9 -/- group n=6). Graphs show aortic aneurysm diameter % increase vs baseline. *P<0.05; **P<0.01 with Welch’s t test. Abbreviations: PPE, Porcine Pancreatic Elastase; Pcsk9, Proprotein convertase, subtilisin/kexin-type 9; AAD, aortic aneurysm diameter

## Discussion

We leveraged clinical and genetic data from 14 cohorts to identify and investigate the genetic determinants of AAA in 39,221 individuals with AAA and 1,086,107 controls. We identified 144 independent AAA-associated variants in 121 loci. We confirmed 24 previously identified AAA genetic risk loci and uncovered 97 novel loci. The increased power in the meta-analysis, compared to the previously published studies, was also evident in the improved performance of PRS derived from the new summary statistics. These data enabled us to prioritize causal genes and pathways for AAA through a combination of functional annotation and gene expression analyses in AAA patients and murine AAA models. We examined the spectrum of phenotypic consequences for AAA risk variants, revealing possible mechanisms through which these variants may lead to disease. Lastly, through a combination of colocalization experiments, as well as analysis in human plasma and *Pcsk9*^-/-^ murine AAA models, we prioritized possible therapeutic targets for the treatment and prevention of AAA.

These findings permit several conclusions. First, this research identifies important fundamental human AAA pathobiology. Our findings highlight not only lipid metabolism, but also vascular development and remodeling, extracellular matrix dysregulation and inflammation as key mechanisms in the pathogenesis of AAA (**Figure 7**). While the dysregulation of these pathways in this disease have been amply demonstrated in published studies of murine models and in diseased human tissues, their identification in the context of population-scale GWAS suggests a role in upstream causation. The putative causal genes identified in this research are in some cases unique to AAA, and in others, already known or suspected to play a role in other cardiovascular diseases. Drug development pipelines and clinical trials are long, expensive, and complex, so new putative interventions need to be carefully chosen. Our findings strongly suggest that treatments that are beneficial for traits that we have now proven to be related to AAA genetically (e.g. atherosclerotic vascular disease and inflammatory conditions) should also be tested for their effect on AAA.

**Figure 7.**
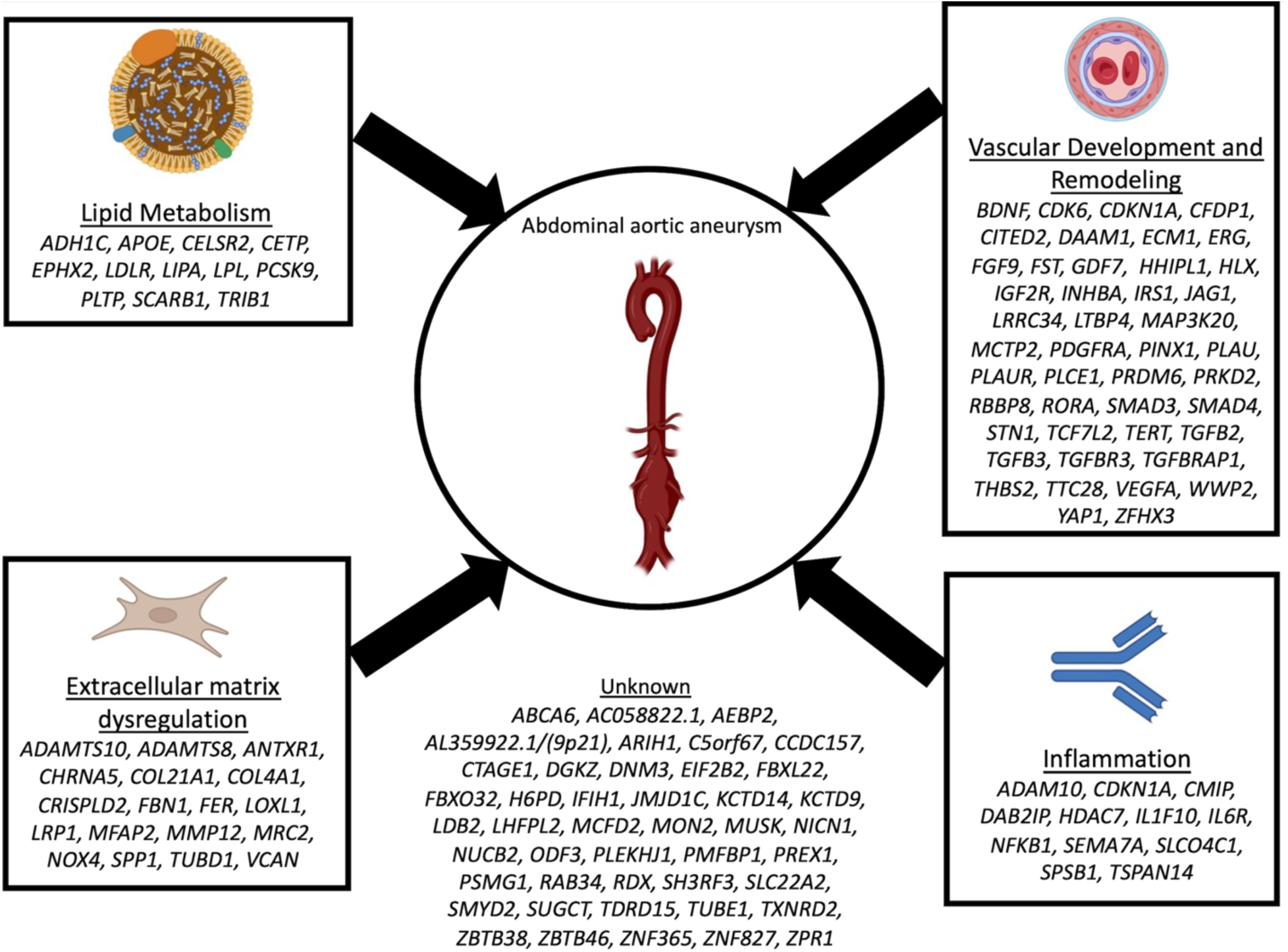
Biological mechanisms underlying genetic loci associated with AAA. The AAA GWAS loci for which we have identified a candidate causal gene are depicted along with the plausible relationship to its underlying biological mechanism. Loci names are based on the candidate causal gene identified in our analysis. However, the biological pathway(s) remain unclear for many associated loci and, as such, the resultant annotation may prove incorrect in some cases.

The genetic results highlight the critical role of lipids and lipid metabolism in AAA pathogenesis. Previous work has demonstrated the likely causal relationship between lipids and AAA (Harrison *et al*., 2018; Klarin *et al*., 2018), and current therapeutic strategies largely focus on LDL-C lowering for broad atherosclerotic risk factor modification in AAA patients rather than specifically for disease prevention or treatment (O’Donnell et al., 2018; Twine and Williams, 2011) in individuals or families at future risk of AAA. Here, we expand upon these findings by observing that 42 of the 121 AAA risk loci are also associated with lipids, supporting the notion of AAA as an end-organ manifestation of atherosclerosis. We additionally prioritize lipid subfractions beyond LDL-C, namely remnant cholesterol, as likely causally related to AAA. Molecules targeting the lipoprotein lipase pathway via *APOC3* (Gaudet et al., 2015) are emerging therapies for cardiovascular disease but remain unstudied in aneurysm patients.

Finally, our results lend human genetic support to *PCSK9* inhibition as a potential therapeutic strategy for AAA. While current data on the effects of statins on aortic expansion are conflicted (Forsdahl et al., 2009; Salata et al., 2018; Twine and Williams, 2011), human genetic evidence has overwhelmingly suggested that LDL cholesterol reduction is likely to reduce AAA risk. Given the dramatic reductions in LDL cholesterol observed with PCSK9i administration (Sabatine *et al*., 2017), assessing whether an LDL cholesterol treatment “floor” exists remains critical. Our genetic analyses suggest a linear dose response for *PCSK9* inhibition on AAA risk –lower LDL cholesterol demonstrates reduced AAA risk regardless of initial levels - and clinical trial evidence demonstrates that LDL cholesterol reduced to values observed in the FOURIER trial (∼30 mg/dL) are safe. In addition, our *Pcsk9* null murine model demonstrated reduced AAA growth following elastase infusion in the absence of a hyperlipidemic background, though small statistical differences in lipid fractions were observed between mice strata. Taken together, these data suggest that the relationship between circulating PCSK9 protein and AAA warrants further investigation.

Our study should be interpreted within the context of its limitations. While this stands as the largest genetic analysis to-date for AAA, the demographics of the cases in our study were overwhelmingly male and European ancestry, thus our ability to detect sex-specific or ancestry-specific genetic associations was limited. Second, our AAA phenotype for many of the cohorts is based on electronic health record data which may have resulted in some misclassification of case status. Such misclassification should, however, reduce statistical power for discovery and, on average, bias results toward the null. Third, for a small number of loci our strategy of gene prioritization identified genes that are not well supported by literature. For example, *CELSR2* or *PLTP* instead of *SORT1* or *MMP9*, respectively. We acknowledge that our gene prioritization scheme is imperfect, as are all current methods for gene prioritization. Fourth, MR methods examining the effects of *PCSK9* inhibition on AAA reflect lifelong exposures to reduced LDL cholesterol levels and may not represent the more acute lipid changes associated with drug administration later in life. Finally, while our analysis suggests that LDL cholesterol reduction through *PCSK9* inhibition is likely to reduce AAA risk, it is unclear whether this treatment is likely to mitigate progression of disease once diagnosed. While the results from our mouse model suggests this to be the case, further investigation into the effects of *PCSK9* inhibition on AAA growth and rupture are warranted.

In summary, we identified 144 independent variants associated with AAA risk, prioritized candidate functional genes at these loci which implicated biological pathways, developed an improved and effective PRS, explored the phenotypic consequences of AAA risk variants through PheWAS, identified candidate causal AAA genes, and multiple lipid pathways and genes that may be targeted for AAA risk reduction, including *PCSK9*. These results are demonstrative of how large-scale analyses of human genetic variation coupled with clinical data can be leveraged and utilized for the treatment of understudied diseases.

## Methods

### Discovery cohorts

#### ARIC

The Atherosclerosis Risk in Communities (ARIC) Study is a population-based cohort recruited from four US communities in North Carolina, Mississippi, Minnesota and Maryland. ARIC identified incident, clinical AAAs by searching hospitalization and death records as well as Medicare data through 2011. Clinical AAA were defined as those who had a hospital discharge diagnosis from any of the above sources, or two Medicare outpatient claims that occurred at least one week apart, with *ICD-9-CM* codes of 441.3 or 441.4, or procedure codes of 38.44 or 39.71, or the following cause of death codes: *ICD-9* 441.3 or 441.4 or *ICD-10* code I71.3 or I71.4 (Folsom et al., 2015; Tang et al., 2016). AAAs based on procedure codes were required to be verified by diagnosis codes. Thoracic, thoracoabdominal, or unspecified aortic aneurysms were treated as non-events. Participants reporting prior AAA surgery or aortic angioplasty at baseline were excluded. Genotyping was performed with Affymetrix Genome-Wide Human SNP array 6.0 and race-specific imputation of variant dosages to the 1000 Genomes Project Phase I version 3 reference panel was performed with IMPUTE2. Association analysis was performed using logistic regression in SNPTEST (Marchini and Howie, 2010) among 408 AAA cases and 8554 non-AAAs of EUR ancestry. Covariate adjustment included age at baseline, sex, pack-years of smoking, and principal components (PC) 1-5.

#### CHB-CVDC and DBDS

Copenhagen Hospital Biobank (CHB) is a hospital driven biobank and includes leftover EDTA blood samples drawn for blood type testing or red cell antibody screening from hospitalized patients in the Danish Capital Region (Sorensen et al., 2021). In addition to genetics, EHR, national socioeconomic and health registries extensively characterize each patient. Patients with AAA are included under the Copenhagen Hospital Biobank Cardiovascular Study (CHB-CVDC) (Laursen et al., 2021). The healthy blood donors from “The Danish Blood Donor Study” (DBDS) are included in the study as controls (Hansen et al., 2019). AAA cases were identified using the following ICD codes (ICD8: 44120/44121/44129; ICD10: I71.3/I71.4). Individuals with aorta dissection were not included in the analysis. AAA cases were compared to the remaining individuals from the DBDS and CHB-CVDC studies excluding individuals with abdominal aortic aneurysm, thoracic aortic aneurysm and intracranial aneurysm using the following ICD codes (ICD8: 44109/44111/44110/44119/44120/44121/44129/44299/43000/43001/43008/43009/43090/43091/43098/43099/43701/43791; ICD10: I71.0/I71.1/I71.2/I71.3/I71.4/I71.5/I71.6/I71.8/I71.9/I72/I60/I67.1). The Infinium Global Screening Array from Illumina was used for genotyping samples from CHB-CVDC and DBDS. Whole-genome sequence data from 8429 Danes along with 7146 samples from North-Western Europe forms a reference panel backbone used for imputation (Helgadottir et al., 2020). The association analysis of 3,079 AAA cases and 180,236 controls was performed with SAIGE (Zhou et al., 2018) using year of birth, sex and 10 PCs as covariates.

#### CHIP+MGI

The Cardiovascular Health Improvement Project (CHIP) is a cohort of individuals treated at Michigan Medicine with linked genotype, EHR, and family history data. The Michigan Genomics Initiative (MGI) is a hospital-based cohort with linked genotype and EHR data from participants recruited during pre-surgical encounters at Michigan Medicine. 534 cases from CHIP were identified as aneurysm in abdominal aorta following diagnosis by Cardiologists and after excluding cases with known dissection. From MGI, 749 cases were identified using ICD codes (ICD9: 441.3/4441.4; ICD10: I71.3/I71.4) after excluding cases with known dissection (ICD9: 441.00-03; ICD10: I71.00-03). After removing samples with related phenotypes (phecodes 440–449.99), a case-control matching strategy was used to identify controls from MGI. Case-control matching was performed using MatchIt package (Ho et al., 2011) in R using birth year, gender, array version and 4 genotype PCs. Samples in CHIP and MGI were genotyped using two versions of customized Illumina Infinium CoreExome-24 bead arrays: UM_HUNT_Biobank_11788091_A1 and UM_HUNT_Biobank_v1-1_20006200_A and imputation. Imputation was performed with the Haplotype Reference Consortium (HRC) panel using Minimac4 (Das et al., 2016). Association analysis of 1,283 AAA cases and 12,202 controls was performed using SAIGE (Zhou *et al*., 2018) with birth year, gender, array version and 4 genotype PCs as covariates.

#### deCODE

Icelandic individuals with AAA were identified from a registry of individuals, using ICD codes (ICD9: 441.3, 441.4, ICD10: I71.3, I71.4) who were admitted at Landspitali University Hospital, in Reykjavik, Iceland, 1980–2016, or diagnosed at private clinic in Reykjavik, Iceland. The Icelandic controls used were selected from individuals who have participated in various GWA studies and who were recruited as part of genetic programs at deCODE. Individuals with known cardiovascular disease were excluded as controls but controls were unscreened for AAA. Genotyping was performed using various Illumina SNP chips and phased using long-range phasing (Kong et al., 2008), followed by imputation of 37.6m high-quality variants from 49,962 whole genome-sequenced Icelanders (Gudbjartsson et al., 2015; Jonsson et al., 2017). The association analysis of 1,656 AAA cases and 265,410 controls was performed using deCODE genetics software (Gudbjartsson *et al*., 2015), and we adjusted for gender, county of origin, current age or age at death (first and second order term included), blood sample availability for the individual, and an indicator function for the overlap of the lifetime of the individual with the time span of phenotype collection. We used LD score regression to account for distribution inflation due to cryptic relatedness and population stratification (Bulik-Sullivan et al., 2015).

#### DiscovEHR

The DiscovEHR cohort is a collaborative effort between Geisinger Health System and the Regeneron Genetics Center. DiscovEHR participants are from Geisinger Health System’s MyCode Community Health Initiative including patients from rural Pennsylvania (USA) recruited from 2007-2021. We included individuals with EHR data, genotype data, and those determined to be of primarily European descent based on SNP-derived principal components analysis. We defined AAA cases based on the presence of at least two instances of any of the following ICD10 codes: 441.3, 441.4, I71.3, I71.4, and required controls to lack any occurrence of the aforementioned ICD10 codes, in addition to the ICD10 codes I71-75, I77-79, K55. We further removed individuals who contributed to the study via the eMERGE consortium. DiscovEHR participants were genotyped on either the Illumina Omni Express Exome or Global Screening Array and imputed from the HRC reference panel using the Michigan Imputation Server. We performed association analysis of 2,238 AAA cases and 105,433 controls using whole genome regression in REGENIE (v1.0), including Age, Age^2^, Sex, Age×Sex, Age^2^×Sex, 10 common variant (MAF>1%) derived principal components, and an array batch indicator as covariates in the model.

#### eMERGE

The electronic Medical Records and Genomics (eMERGE, Phase 3) network is a consortium with EHR and genetic data on ∼100,000 patients from 12 institutions across the US, including ∼20% samples from African ancestry. Samples with 2 or more instances of ICD9 codes 441.3/4441.4 or ICD10: I71.3/I71.4 were defined as cases and samples without instance of case ICD codes were defined as controls. Case control matching was performed separately on EUR and AFR samples to retain 5 controls per each case matching based on age and sex using MatchIt R program. Samples in eMERGE Phase III were genotyped on 78 different Illumina and Affymetrix SNP array platforms. Data from different sites and genotyping platforms were imputed using the HRC reference panel on Michigan Imputation Server and then merged into one set (Macrae et al., 1989; Stanaway et al., 2019). We identified 3092 cases and 15,025 controls in the EUR dataset and 119 cases and 603 controls in the AFR dataset, and logistic regression was performed on unrelated samples within ancestry groups using PLINK 2.0. All models were adjusted by clinical site and first 5 principal components.

#### HUNT

The Trøndelag Health (HUNT) Study (Krokstad et al., 2013) is a population-based health study from the Trøndelag region in Norway. AAA cases were defined by ICD-9 codes 441.3 and 441.4 and ICD-10 codes I71.3 and I71.4. HUNT samples were genotyped using the Illumina HumanCoreExome array and imputed with Minimac3 (Das *et al*., 2016) using a combined reference panel consisting of the HRC v1.1 combined with whole genome sequencing of 2,201 HUNT participants. Association testing of 734 cases and 68,901 controls was performed using SAIGE (Zhou *et al*., 2018) with batch, sex, birth year, and PC1-4 as covariates.

#### Mayo VDB

Mayo Vascular Disease Biorepository (VDB) at the Mayo Clinic was established to archive DNA, plasma, and serum from patients with suspected atherosclerotic cardiovascular disease (ASCVD) referred for noninvasive vascular evaluation and exercise stress testing at the Mayo Clinic Gonda Vascular Center from January 14, 2006 to July 24, 2020. After exclusion of overlapping samples between Mayo VDB Dataset and eMERGE III V3 Imputed Array Dataset (Stanaway *et al*., 2019), AAA cases were defined as having an infrarenal abdominal aortic diameter ≥3 cm or a history of open or endovascular AAA repair based on an electronic phenotyping algorithm using Natural Language Processing on ultrasound reports, validated by manual chart review. Controls were not known to have AAA and had no ICD-9 diagnosis codes for AAA. Genotyping was performed using three different Illumina platforms, including Illumina Human660W-Quad V1, HumanCoreExome Beadchip, and Human 610 Quad V1 platforms. We combined samples genotyped on different platforms by genotype imputation based on a HRC v1.1 panel. Association testing of 771 AAA cases and 4,913 controls was performed on the imputed dataset using SAIGE software with the following covariates: study enrollment age, sex, genotyping platform, first 5 principal components of ancestry.

#### MVP

In the Million Veteran Program (MVP), individuals aged 18 to over 100 years have been recruited from 63 VA Medical Centers across the United States. From the participants passing quality control in MVP, individuals were defined as having AAA or being a disease-free control using a previously utilized (Klarin *et al*., 2020) definition initially proposed by Denny et al (Denny et al., 2013). AAA cases were defined as the presence of two instances of any of the following ICD-9/10 codes in a participant’s EHR: 441.3, 441.4, I71.3, I71.4. Controls were defined as possessing zero occurrences of the aforementioned ICD codes, as well as zero occurrences of the ICD-9 codes 440-448, or ICD-10 codes I71-75, I77-79, K55. In our MVP analysis, we evaluated 17,672 AAA cases and 303,695 controls of European ancestry, and 1,888 AAA cases and 87,728 controls of African ancestry. Genotyped and imputed DNA sequence variants in individuals of were tested for association with AAA using logistic regression adjusting for age, sex, and 5 principal components of ancestry assuming an additive model using the PLINK2.0 statistical software program.

#### NZ AAA Genetics Study

The Vascular Research Consortium of New Zealand recruited New Zealand men and women with a proven history of AAA (infra-renal aortic diameter ≥ 30 mm proven on ultrasound or CT scan). Approximately 80% had undergone surgical AAA repair (typically AAA’s > 50-55 mm in diameter). The vast majority of cases (>97%) were of Anglo-European ancestry. The control group underwent an abdominal ultrasound scan to exclude (>25 mm) concurrent AAA and Anglo-European ancestry was required for inclusion. Controls were also screened for peripheral artery disease (PAD; using ankle brachial index), carotid artery disease (ultrasound) and other cardiovascular risk factors. All participants were genotyped in two separate case-control cohorts using the Affymetrix SNP6 (cohort 1; 608 AAA cases and 612 controls) or Illumina Omni2.5 (cohort 2; 397 cases and 384 controls) GeneChip arrays and had call rates >95% (mean 99.2%). Imputation was then conducted using IMPUTE 2.2 run on the BCISNPmax database platform (version 3.5, BCI Platforms, Espoo, Finland). The reference haplotypes were based on the 1000 Genomes June 2011 release. Imputed calls were ?ltered by quality score (>0.9) to restrict to higher quality imputed SNPs. The genomic in?ation factors were 1.06 and 1.05 respectively (MAF >0.05, 5.4 million SNPs). Case-control genome wide association analyses were conducted using PLINK (version 1.07).

#### PMBB

Penn Medicine biobank (PMBB) recruits patients from throughout the University of Pennsylvania Health System for genomic and precision medicine research. Participants actively consent to allow the linkage of biospecimens to their longitudinal EHR. Currently, >60 000 participants are enrolled in the PMBB. A subset of ∼45000 individuals who have undergone whole exome sequencing and genotyping, performed through a collaboration with the Regeneron Genetics Center. A further subset of ∼23000 subjects with imputed genotype data was used in this analysis. 19,515 recruits were genotyped on three different arrays, Illumnia Quad Omni, Global Screening Assay v1 and Global Screening Assay v2. Genotype imputations were performed using the Eagle2 (Loh et al., 2016) and Minimac softwares (Das *et al*., 2016) on the Michigan Imputation server and were completed for all autosomes with the HRC reference panel. AAA cases were found using the AAA phecode definition of 442.11, meaning at least 1 encounter with abdominal aortic aneurysm diagnosis codes and excluding other confirmed disease of the arteries. Controls were defined as all others with genotype data and no evidence of AAA. GWAS of 388 cases and 9879 controls was performed using plink software with age at recruitment, gender, genetic determined ancestry and the first 10 principal components.

#### TABS

The Triple A Barcelona Study (TABS) is a hospital-based study recruiting individuals with AAA treated in the Hospital de la Santa Creu i Sant Pau in Barcelona, Spain. All individuals have repetitive measurements of the abdominal aortic diameter, either by CT-scan or by ultrasound along with anthropometric and clinical information. Cases were defined as those with dilation of the abdominal aorta with a diameter higher than 30 millimeters. Type B dissections that progress to AAA, saccular aneurysms or thoracic aneurysms were excluded. In addition to 42 cases from TABS, 10 cases and 82 controls were leveraged from the Triple A Genetic Study (TAGA) (Peypoch et al., 2020). Also, 401 controls were leveraged from the RETROVE study (Vazquez-Santiago et al., 2017). Genome-wide genotyping was performed using the Infinium Global Screening Array-24 v2.0 from Illumina (coverage 665,608 variants) and imputed to the HRC reference panel using the Michigan server. Association analysis of 52 AAA cases and 483 controls was performed using SAIGE (Zhou *et al*., 2018) with birth year, sex, batch and PCs as covariates.

#### UKAGS+VIVA and UKBB

The UK Aneurysm Growth Study (UKAGS), The Viborg Vascular (VIVA) and UK Biobank (UKBB) cohorts are described together since cases from all three cohorts were compared with controls from UKBB. The UK Aneurysm Growth Study (UKAGS) is a prospective study of men attending the NHS aneurysm screening programmes in the UK. Men recruited into the UKAGS completed a postal questionnaire to obtain information on smoking, comorbidities and medications. Screening outcomes (ultrasound measured AAA diameter) were obtained directly from the AAA screening programmes. The VIVA screening trial is a randomized, clinically controlled study designed to evaluate the benefits of vascular screening and modern vascular prophylaxis in a population of 50,000 men aged 65-74 years, randomized to either receive an invitation for vascular screening or being a control. Controls were selected from the UK biobank (UKBB), a large prospective study with over 500,000 participants aged 40–69 years when recruited between 2006–2010. To identify AAA cases in UKBB the following ICD and OPCS codes were searched for in the UKBB hospital inpatient data. Any individual with one or more of the following codes was defined as a case: ICD9: 441.3, 441.4; ICD10: I71.3, I71.4; OPCS4: L184, L185, L186, L194, L195, L196, L271, L275, L276, L281, L285, L286. Controls were selected from UKBB so as to exclude individuals with any potential aortic pathology such as thoracic aortic aneurysm using the following ICD codes as exclusion criteria: ICD9: 441.00, 441.01, 441.02, 441.03, 441.1, 441.2, 441.5, 441.6, 441.7, 441.9; ICD10: I71.0, I71.1, I71.2, I71.5, I71.6, I71.8, I71.9. The group of controls was then restricted to the UKBB ‘in white British ancestry subset’ and age and sex matched control groups were selected (without overlap) for each of the separate case groups used in the discovery and validation cohorts (see Additional validation cohorts for PRS). All participants in UKAGS and VIVA were genotyped using the UKBB Axiom Array. UKBB participants were genotyped using the UKBB Axiom array and UKBB BiLEVE Axiom array. All imputation, including for UKBB was carried out on the Michigan Imputation Server, Minimac 4, 1000G Phase3 v5 (GRCh37/hg19) reference panel (Rsq filter = 0.3). Association analysis of 3,595 AAA cases (3,209 UKAGS + 386 VIVA) and 15,773 controls (UKBB) was performed with PLINK v2.00a, with following parameters, --geno 0.02, --hwe 1e-8, --maf 0.01. In addition, a non-overlapping set of UKBB subjects was used for association analysis of 1,241 AAA cases (1081 males, 160 females) and 6,276 controls (5466 males, 810 females) with SNPTEST v2.5.2, with following parameters, -frequentist additive, -method expected.

All research participants provided informed consent and local IRB approval was obtained. See supplementary materials for additional details.

#### QC prior to meta-analysis

A central level QC was performed on summary statistics from discovery cohorts. QQ plots (**Figure S1**) and GC lambda were calculated for each cohort (**Table S1**). Cohorts that reported variants in hg38 version were liftover to hg19 version. Cohorts that reported odds ratios were converted to beta/effect estimate using log (odds ratio). We checked consistency of effect estimates in 9 index variants that were reported in Jones et al. (Jones *et al*., 2017). While we observed some heterogeneity, the effect estimates were generally consistent among well powered studies (**Figure S2**). Variants with minor allele count of 3 and imputation R^2^ < 0.3 were excluded from individual summary statistics.

#### Meta-analysis and internal replication

Meta-analysis of 17 discovery cohort summary statistics was performed by METAL (Willer et al., 2010) in standard error mode with genomic control on. After meta-analysis, variants that were present in only one cohort were excluded from downstream analysis. For internal replication, another meta-analysis summary statistics were generated in the same approach, only without the MVP (EUR) cohort.

#### Definition of loci

Independent loci were defined as variants >1 Mb and >0.25 cM apart with at least one genetic variant associated with AAA at a genome-wide significance threshold of P < 5 × 10^−8^. Index variants are the variants with lowest association p-value in every locus.

#### Conditional analysis

To perform conditional analysis, we first defined the loci as +/- 1 Mb from each of the 121 index SNPs. GCTA COJO (Yang et al., 2011) and specifically the “cojo-cond” function was performed iteratively using a reference panel created from individuals in Penn Medicine BioBank that represented the demographics of AAAgen (17:1 EUR to AFR ancestry). Iterations were performed at each locus until a minimum number of independently, genome-wide significant SNPs were identified. The maximum number of iterations performed was 4 at the rs10455872 locus.

#### Effect estimates for polygenic risk score

To calculate PRS, we used PRScs (Ge *et al*., 2019) that calculates posterior SNP effect estimates from original GWAS effect estimates using a Bayesian approach. PRScs uses information from an external LD reference panel for this calculation. PRScs calculated posterior SNP effect sizes using a pre-computed LD reference panel from EUR ancestry (ldblk_1kg_eur.tar.gz). These effect estimates were used to calculate AAA PRS of individuals in validation cohorts using PLINK2 --score command (Purcell et al., 2007).

#### Additional validation cohorts for PRS

Additional AAA case cohorts from Oxford (UK), Uppsala (Sweden) and Utrecht (Netherlands) became available during the conduct of the study. Those from Oxford and Uppsala were genotyped alongside remaining cases from the UKAGS and VIVA study that had not been included in the discovery study and were analyzed together in comparison to controls taken from UK Biobank. Finally, the Aneurysm Consortium AAA GWAS (Bown *et al*., 2011) had not met QC criteria for inclusion in the discovery analysis but was available for validation of polygenic risk scores.

#### OxAAA

Multimodal Assessment of Aortic Aneurysm Disease Pathogenesis: Oxford Abdominal Aortic Aneurysm Study (OxAAA). Single centre study at the Oxford University Hospitals NHS Trust, funded by the NIHR Oxford Biomedical Research Center. IRAS ID: 122591. REC name: South Central - Oxford C Research Ethics Committee. REC reference: 13/SC/0250.

#### UppsalaAAA

Uppsala Abdominal Aortic Aneurysm Cohort Study, part of the collection Swedish Cohort Consortium (Cohorts.se). A prospective population-based case-control study which aims to find biomarkers for the cause and progression of AAA and investigate how persons with or without an AAA experience their health. All 65-year-old men, identified through the National Population Registry, were invited to an ultrasound examination. Financial support was provided by the Swedish Research Council (grant 2012-1978), the Swedish Heart and Lung Foundation, the King Gustaf V’s and Queen Victoria’s Freemason Foundation, and Regional Uppsala-Örebro Research Grant

#### AC

Aneurysm Consortium (AC), cases of AAA together with available DNA recruited in eight centers in the UK, Australia, and New Zealand and also samples from the United Kingdom Small Aneurysm Trial from inpatient populations, outpatient clinics, or population screening programs in the participating centers. AAA ascertainment (infrarenal aortic diameter of >30 mm) was by either ultrasonography or by cross-sectional imaging except for patients who presented with acute rupture and for whom it was assumed that the AAA was >55 mm. Further details, including funding, ethical approval in Bown et al. (Bown *et al*., 2011).

#### WTCCC2

Wellcome Trust Case Control Consortium 2 data used included samples from the 1958 British Birth Cohort and from the UK National Blood Service. Further details in Bown et al. (Bown *et al*., 2011) and https://www.wtccc.org.uk/.

#### PRS validation set 1

This set includes 808 cases from UKAGS and 4 cases from VIVA that were not used in meta-analysis, 247 cases from UppsalaAAA, 71 cases from OxAAA, and 5810 controls from UKBB. All participants in UKAGS, UppsalaAAA, OxAAA and VIVA were genotyped using the UK Biobank Axiom Array. UKBB participants were genotyped using the UK Biobank Axiom array and UK Biobank BiLEVE Axiom array. The following QC filters were used: 1) White British ancestry only, 2) 1 of any pairwise kinships removed, 3) outliers identified by PCA and excluded, 4) batch missing test (--test-missing in PLINK) to exclude variants with highly significant difference in missingness between cases and controls (P <0.001), 5) variants with missing call rates > 2% excluded. A pre-imputation check using the McCarthy Group tool was performed. All imputation, including for UKBB was carried out on the Michigan Imputation Server, Minimac 4, 1000G Phase3 v5 (GRCh37/hg19) reference panel, Rsq filter = 0.3.

#### PRS validation set 2

This set includes 1887 cases from AC and 5437 controls from WTCCC. The case and control cohorts were separately genotyped with the Illumina 670K BeadChips. Raw intensity data were normalized with BeadStudio, and genotypes were called concurrently from the combined case control data set with the Illuminus algorithm. The following QC filters were used: 1) Identity-by-state (IBS) clustering carried out in PLINK and extreme outlier individuals excluded, 2) Batch missing test (--test-missing in PLINK) used to exclude variants with highly significant difference in missingness between cases and controls (P <0.001), 3) variants with missing call rates > 2% excluded. A pre-imputation check using the McCarthy Group tool was performed. Imputation was carried out on the Michigan Imputation Server, Minimac 4, 1000G Phase3 v5 (GRCh37/hg19) reference panel, Rsq filter = 0.3.

#### Cox Proportional Hazards model

We performed another meta-analysis without UKBB by METAL (Willer *et al*., 2010), followed by calculation of PRS weights by PRScs (Ge *et al*., 2019), as described above. The UKBB hospital registry data and cause of death data up to March 2020, was used to test the predictive performance of the AAAgen PRS using Cox Proportional Hazards models. The PRS was adjusted with four first PCs and inverse normalized for the analysis. Analysis only included the white British subset due to the low number of AAA cases in the other ancestries. The baseline of the model was set to the individuals’ clinical assessment date. First occurrence of ICD9 codes 441.3 and 441.4, or ICD10 codes I71.3 and I71.4 was recorded together with the date of diagnosis for all AAA cases. 10-year survival was modeled with follow-up time as the time-scale. Prevalent cases (N=213) were excluded from the analyses and individuals that deceased during the follow-up for other causes than AAA were censored. The final analysis included 838 incident AAA cases and 329,983 non-cases with median follow-up time of 5.04 and 10.0, respectively.

#### Gene-set enrichment

We performed gene-set enrichment analysis using DEPICT (Pers *et al*., 2015). DEPICT uses reconstituted gene sets, consisting of 14,462 gene sets obtained from multiple sources and reconstituted using 77,840 publicly available microarray expression datasets. In the reconstituted gene sets every gene in the human genome is assigned a z-score for membership in the set. Using genes from GWAS loci, DEPICT calculates an enrichment p-value for reconstituted gene sets. We used AAA summary statistics and clumping with a threshold of 5 × 10^−8^ as input to DEPICT. Genes with z-score > 2.58 (p-value < 0.01) were considered as members of gene sets. The overlap between the gene sets were calculated using Jaccard index. In **Figure 2a**, we highlighted the top 30 DEPICT reconstituted gene sets excluding terms with “PPI subnetwork” and “positive regulation” in presence of “regulation”.

#### Tissue enrichment

Calculation of enrichment in specific tissue types were calculated by stratified LD score regression (LDSC) (Finucane *et al*., 2018), a method for partitioning heritability. Using GWAS summary statistics and tissue specific chromatin marks this method calculates the enrichment of per-SNP heritability. First, hapmap3 variants with MAF> 0.01 were obtained from the AAA summary statistics using munge_sumstats.py. Then, the analysis was performed using the –-h2-cts option in LDSC. We reported p-values of enrichment of per-SNP heritability in various chromatin marks from ENCODE and Roadmap Epigenome.

#### Cell-type enrichment

Processing of single cell RNA (scRNA) sequencing data from aorta is described in Davis et al. (Davis *et al*., 2021). Calculation of enrichment in specific cell types using scRNAseq data was performed using RolyPoly (Calderon *et al*., 2017). RolyPoly uses a regression-based polygenic model to prioritize relevant cell types from GWAS summary statistics and scRNAseq data. Average expression per cell type was obtained by using Seurat (Satija et al., 2015) AverageExpression() function. This generates a matrix with genes as rows and cell types as columns. Columns were normalized using normalize.quantiles() and rows were normalized using scale() function in R. Since RolyPoly does not work with negative values, we used the absolute values for each entry. To link gene expression with GWAS variants, we used block annotation of 25 kb around the gene’s start. For LD statistics we used 1000 genome phase 3 data pre-computed by RolyPoly. We ran the function rolypoly_roll() with 200 bootstrap iterations. P-values from the bootstrap analyses were reported.

#### Identification of coding variants

First, we identified variants that are in LD (>0.8) with index variants using 42,119 unrelated EUR individuals from CHIP+MGI. Variant effect predictor (VEP) (McLaren et al., 2016) was used to predict molecular consequences of these variants. We reported genes with at least one variant altering amino acid (missense variant) from VEP output. We did not observe any gene with stop-gained or frameshift variants.

#### Genes causing related monogenic phenotypes

A list of 53 genes curated in Renard et al. (Renard *et al*., 2018) that potentially causes monogenic form of heritable thoracic aortic aneurysm and dissection was incorporated. For a range of related/risk factor phenotypes and disorders (**Table S9**), we obtained ClinVar database (Landrum et al., 2018) entries with pathogenic or likely pathogenic variants on 23^rd^ June, 2021. From these entries, genes (either start or end) that are within 1 Mb of index variant were incorporated.

#### PoPS for gene prioritization

We used PoPS (Weeks *et al*., 2020), a similarity-based gene prioritization method that uses a large set of publicly available RNA sequencing, curated pathway annotation, and predicted protein-protein interaction datasets. First, PoPS calculates gene level association statistics from GWAS variant level association statistics and MAGMA gene annotations. Second, Features are selected based on pre-computed statistics from public resources. Finally, PoPS computes a score for each gene. Based on these scores, for each genome-wide significant locus, we ranked the genes within 1 Mb (either direction) of the index variant and reported the gene with the highest score as the gene prioritized by PoPS.

#### eQTL Colocalization

We performed colocalization analyses using the R package coloc (Giambartolomei et al., 2014). Coloc performs an approximate Bayes factor analysis with association statistics. The function coloc.abf() was used to calculate the posterior probabilities for: (H0) no association with either trait; (H1/H2) association with one of the two tested traits; (H3) association for both traits but different causal variants; and (H4) association for both traits with the same causal variant. A high posterior probability for H4 (PP4) indicates colocalization of the two trait associations. Colocalization was performed with eQTLs in aorta, liver, whole blood, adipose subcutaneous and visceral omentum from GTEx v8. Variants within 500 kb in either direction of GWAS index variants were extracted for the analyses. A PP4 threshold of 0.5 was used to report colocalization between GWAS and eQTL locus.

#### Transcriptome wide association study

We used the paradigm of transcriptome wide association study (TWAS) that performs gene-based association tests. These methods are used to test the association between gene expression predicted by cis-eQTLs and phenotype. The MetaXcan package (Barbeira et al., 2018) was used to run TWAS with AAA summary statistics. Briefly, we used GWAS tools from the MetaXcan package for summary statistics harmonization and imputation. The imputation step imputes missing GWAS variants using present GWAS variants and the GTEx genotypes. Next, we ran SPrediXcan with the imputed variants and the MASHR expression model (eQTL) of aorta, liver, whole blood, adipose subcutaneous and visceral omentum from GTEx v8 (Consortium, 2020). For each tissue, significance threshold was decided with correction for multiple testing (0.05/number of genes).

#### Bulk RNA sequencing from AAA patients

Abdominal aortic tissue was surgically resected during open repair for an abdominal aortic aneurysm. Samples were placed in a sterile field, partitioned into 0.5 cm x 0.5 cm pieces, placed in RNAlater (Qiagen GmbH, Hilden, Germany) and stored at room temperature between 1-3 days. The RNAlater solution was removed, and the tissue was stored at -80 °C. Tissues were cryopulverized using a CP02 instrument (Covaris), and an aliquot of the powdered tissue was used for isolation of total RNA using Trizol (Invitrogen) and RNeasy Mini Kit (Qiagen, GmbH, Hilden, Germany). Briefly, the RNA in the aqueous phase from the Trizol extraction was transferred to a new tube and mixed with an equal volume of 70% ethanol before processing on an RNEasy Mini kit column according to the manufacturer’s instructions. RNA was eluted in 50ul of RNAse-free water. Concentrations were measured using the Qubit RNA Broad Range kit (Thermo Scientific, USA), followed by RNA integrity (RIN) evaluation by RNA TapeScreen on the Agilent 2200 TapeStation (Agilent Technologies, USA). RNA samples with a RIN > 7 were used in RNA sequencing analyses. Samples were prepared for sequencing using the Takara SMART-Seq v4 Ultra Low Input RNA Kit plus Nextera XT. Samples (n=126) underwent paired-end sequencing by MedGenome. FASTP (Chen et al., 2018) was used for adaptor trimming of the paired end reads. FASTQC was used for quality control. Reads were aligned to hg38 with annotation from GENCODE Human genome release 39 using STAR (Dobin et al., 2013). Samples were not included in downstream analysis if less than 70% STAR alignment, less than twenty thousand reads, or if the FASTQC GC content curve deviated significantly from a normal distribution centered around 50%. 15 abdominal aortic aneurysm RNA samples passed quality control. FeatureCounts (Liao et al., 2014) was used to aggregate a gene counts matrix. DESeq2 (Love et al., 2014) was used for variance stabilization transformation (VST) normalization. To determine if a gene was expressed or not, a VST expression threshold of 6 was used. This cutoff was chosen based on a density plot of all transcript expression that yielded a bimodal distribution (**Figure S12**). A cutoff of 6 effectively eliminated genes with no expression.

#### Gene expression by qPCR

The study was approved by the Ethical Committee of *Investigación Clínica del Hospital Santa Creu i Sant Pau*. Written informed consent was obtained from all patients. The study conformed to the principles outlined in the Declaration of Helsinki. All patients underwent surgery at Hospital de la Santa Creu i Sant Pau (HSCSP). Samples were obtained from the remaining mid-infrarenal aortic wall after exclusion and prosthetic replacement of AAA. Normal aortas (NA) were obtained from healthy aorta from multi organ donors and samples were also taken from the mid-portion of the infrarenal abdominal aorta at organ harvest. When a luminal thrombus was present it was separated before the aorta biopsy was taken and aortic tissue was washed twice with cold phosphate buffered saline (PBS). A portion of each sample was placed in a RNAlater solution (Qiagen GmbH, Hilden, Germany) and stored at 4°C for 24 hours before long-term storage at -80°C until further processing for RNA isolation. Further information can be found in (Sola-Villa *et al*., 2015). Tissues were homogenized in the FastPrep-24 homogenizer and Lysing Matrix D tubes (MP Biomedicals, Solon, OH, USA). RNA was extracted using Trizol (Invitrogen, Carlsbad, CA, USA) following the manufacturer’s instructions. cDNA was prepared by reverse transcribing 1 µg RNA with a High-Capacity cDNA Archive Kit with random hexamers (Applied Biosystems, Foster City, CA, USA). mRNA expression of the selected genes was studied by real-time PCR in an ABI Prism 7900HT using pre-designed validated assays (TaqMan Gene Expression Assays; Applied Biosystems) and universal thermal cycling parameters. Relative expression was expressed as transcript/β-actin ratios.

#### PheWAS analysis

We performed a PheWAS of 121 independent variants against the entirety of the MRC-IEU open GWAS project (Elsworth *et al*., 2020). The full summary statistics were then filtered to exclude the following phenotypes: mQTLs, pQTLs, eQTLs, sex specific GWAS and non-EUR and non-mixed ancestry GWAS. After calculating the node and edge maps using NETMAGE (Sriram et al., 2020), only connections that represent a weight of >3 SNPs were used to make the graph more interpretable. Raw network statistics were calculated in Gephi (Bastian et al., 2009) including modularity or cluster ID using Blondel et al. (Blondel et al., 2008). This method works to create modules that minimize the number of edges that enter and leave the module. The granularity of the definition of the modules or resolution can be adjusted, with lower resolution generating a higher number of modules. Our optimization efforts lead us to set the resolution parameter as 0.561 generating 7 distinct modules. Colocalization analysis with the largest available GWAS of lipid (Graham *et al*., 2021), blood pressure (Evangelou *et al*., 2018) and smoking (Liu *et al*., 2019) traits were performed by coloc (Giambartolomei *et al*., 2014), as described in the eQTL colocalization section.

#### Mendelian randomization

Genetic associations between lipoprotein fractions (exposure) and AAA outcome were tested using initially using inverse-variance weighted MR for a single lipid exposure, and then using the MR-BMA methodology for multivariable models (Zuber *et al*., 2020). MR-BMA is an extension of multivariable MR utilizing a Bayesian variable selection method in an effort to identify likely causal risk factors among correlated exposures. In the primary analysis, the instrumental variables consisted of independent genetic variants (r^2^ < 0.001 based on 1000 Genomes (Genomes Project et al., 2015) European ancestry Reference Panel) associated with any major lipoprotein at genome-wide significance in the UK Biobank based on 361 194 European-ancestry participants as previously described (Levin *et al*., 2021). Genetic associations with circulating levels of major lipoprotein-related traits in blood (ApoB, LDL-C, HDL-C, TG, ApoA1, and non-HDL-C were used as exposures in two models: one containing LDL-C, HDL-C, ApoA1, ApoB, and triglycerides (n = 519 independent genetic variants), and another substituting non-HDL-C for LDL-C (n = 450 independent genetic variants) (**Figure 5a**). The subsequent MR-BMA analysis was completed using AAA GWAS summary statistics from the current study. Variable selection was based on marginal inclusion probabilities for which an empirical permutation procedure was used to derive P values. The Nyholt procedure of effective tests was used to account for the strong correlation among the lipoproteins, with a multiple testing–adjusted P value of P=0.05 set as the significance threshold (Nyholt, 2004). Further details of statistical analysis are described in the supplementary methods.

#### Proteome-wide Mendelian Randomization

To identify possible therapeutic targets for AAA, we performed a proteome-wide MR analysis using high-confidence cis-acting genomic instruments for circulating plasma proteins passing pleiotropy and consistency filters as previously described (Zheng *et al*., 2020). In brief, variants were selected that were associated with any protein at genome-wide significance (P < 5×10^−8^). For this analysis, we focused only on *cis-*pQTLs previously classified as having the highest relative level of reliability (“Tier 1”). After excluding the major histocompatibility complex region, LD clumping to identify instruments composed of independent pQTLs was performed using the TwoSampleMR package (Hemani et al., 2018) with an r^2^ threshold of < 0.001. After restricting to 717 circulating proteins with overlapping proteomic and genomic data (**Table S26**, we performed a pQTL screen: 1) for exposure-outcome pairs with 2 or more available genetic instruments as proxies for the exposure, inverse variance-weighted MR was performed; 2) when only a single genetic proxy for the exposure was present, Wald-ratio MR was performed. We set an Benjamini-Hochberg FDR P < 0.05 for statistical significance. For each protein-AAA association in cis-pQTL-MR above, we performed colocalization to provide supporting evidence for causal assocations among traits. Colocalization was performed using HyPrColoc (Foley et al., 2021).

#### *Pcsk9* ^*-/-*^ mice and AAA progression

All animal protocols were approved by the Administrative Panel on Laboratory Animal Care at Stanford University (http://labanimals.stanford.edu/) and the VA Palo Alto Health Care System Institutional Animal Care and Use Committee and followed the National Institutes of Health and U.S. Department of Agriculture Guidelines for Care and Use of Animals in Research. Male mice (wild type and *Pcsk9*^*-/-*^ mice on a C57BL/6J background) were purchased from The Jackson Laboratory.

#### PPE infusion model

The PPE infusion model to induce mouse AAA was performed as previously described at 10 weeks of age (Sabatine *et al*., 2017). The proximal and distal aorta was temporarily ligated or clamped, followed by an aortotomy above the iliac bifurcation. A catheter was used to infuse the aorta for 5 minutes at 120 mmHg with saline containing type I porcine pancreatic elastase (2.5U/mL; Sigma Aldrich), and the aortotomy was then repaired. The induced AAA aortic segment (between the left renal artery and the bifurcation) was harvested at 28-days post-surgery.

#### Lipid measurements

Plasma cholesterol, LDL, and HDL levels were measured at Stanford Animal Diagnostics Laboratory on the Siemens Dimension EXL200/LOCI analyzer.

#### Aortic diameter measurements by ultrasound imaging

At baseline and 3, 7, 14, 21, and 28 days after aneurysm induction, B-mode ultrasound imaging was performed on the operated mice to assess the AAD as previously described (Sabatine *et al*., 2017).

#### Murine AAA Model Statistical Analysis

Data are presented as means ± SEM. Groups were compared with Welch’s t-test for parametric and Mann-Whitney for non-parametric data. Shapiro Wilk test was performed to test normality. A value of P < 0.05 was considered statistically significant.

## Data Availability

All data produced in the present study are available upon reasonable request to the authors

## Funding/Cohort-Specific Acknowledgements

1. Greg Jones - NZ Vascular Research Consortium AAA Cohort
  a. Health Research Council of New Zealand (14/155, 17/402, 20/144)
2. Corry Gellatly - UK Aneurysm Growth Study (UKAGS)
  a. CG is funded by British Heart Foundation grants CS/14/2/30841 and RG/18/10/33842.
3. Matthew Bown - UK Aneurysm Growth Study (UKAGS)
  a. UKAGS is funded by British Heart Foundation grants CS/14/2/30841 and RG/18/10/33842.
4. Katie Saxby - UK Aneurysm Growth Study (UKAGS)
  a. Wellcome Trust Doctoral Training Programme reference 222959/Z/21/Z
5. Shefali S. Verma, Todd L. Edwards, Marylyn D. Ritchie - PMBB, eMERGE
  a. eMERGE Network (Phase III): This phase of the eMERGE Network was initiated and funded by the NHGRI through the following grants: U01HG8657 (Group Health Cooperative/University of Washington); U01HG8685 (Brigham and Women’s Hospital); U01HG8672 (Vanderbilt University Medical Center); U01HG8666 (Cincinnati Children’s Hospital Medical Center); U01HG6379 (Mayo Clinic); U01HG8679 (Geisinger Clinic); U01HG8680 (Columbia University Health Sciences); U01HG8684 (Children’s Hospital of Philadelphia); U01HG8673 (Northwestern University); U01HG8701 (Vanderbilt University Medical Center serving as the Coordinating Center); U01HG8676 (Partners Healthcare/Broad Institute); and U01HG8664 (Baylor College of Medicine). We would also like to acknowledge the following eMERGE members who contributed to the eMERGE data for this manuscript: D. Crosslin, J. Denny, M. Palmer, SA. Pendergrass, and I. Stanaway.
6. Ozan Dikilitas - Mayo, eMERGE
  a. OD is supported by Mayo Clinic Clinician Investigator training program.
7. Ben Brumpton - HUNT
  a. The Trøndelag Health Study (The HUNT Study) is a collaboration between HUNT Research Center (Faculty of Medicine and Health Sciences, NTNU, Norwegian University of Science and Technology), Trøndelag County Council, Central Norway Regional Health Authority, and the Norwegian Institute of Public Health. The genotyping in HUNT was financed by the National Institutes of Health; University of Michigan; the Research Council of Norway; the Liaison Committee for Education, Research and Innovation in Central Norway; and the Joint Research Committee between St Olav’s hospital and the Faculty of Medicine and Health Sciences, NTNU.
8. Sander W. van der Laan
  a. SWvdL is is funded through EU H2020 TO_AITION (grant number: 848146).
  b. SWvdL has received Roche funding for unrelated work.
  c. We are thankful for the support of the Netherlands CardioVascular Research Initiative of the Netherlands Heart Foundation (CVON 2011/B019 and CVON 2017-20: Generating the best evidence-based pharmaceutical targets for atherosclerosis [GENIUS I&II]), the ERA-CVD program ‘druggable-MI-targets’ (grant number: 01KL1802), and the Leducq Fondation ‘PlaqOmics’.
9. Maria Sabater-Lleal - TABS/AAA expression Sant Pau
  a. TABS is funded by grant PID2019-109844RB-I00 from the Spanish Ministry of Science and Innovation. The genotyping service was carried out at CEGEN-PRB3-ISCIII and supported by grant PT17/0019, of the PE I+D+i 2013-2016, funded by ISCIII and ERDF. Maria Sabater-Lleal is supported by a Miguel Servet contract from the ISCIII Spanish Health Institute (CP17/00142) and co-financed by the European Social Fund.
10. Gregory T. Jones - NZ Vascular Research Consortium AAA Cohort
  a. Health Research Council of New Zealand (14/155, 17/402, 20/144)
11. Cristen Willer - CHIP/MGI, HUNT
  a. CJW is supported by NIH grants R35-HL135824-03 and R01-HL142023-02
12. Scott Damrauer - MVP, PMBB
  a. SMD is supported by IK2-CX001780. This publication does not represent the views of the Department of Veterans Affairs or the United States Government.
13. Karina Banasik, Alex Christensen & Søren Brunak - CHB/DBDS
  a. KB and SB acknowledge the Novo Nordisk Foundation (grants NNF17OC0027594 and NNF14CC0001)
  b. AHC is supported by The Independent Research Fund Denmark (0134-00363B) and The Novo Nordisk Foundation (NNF20OC0065799)
14. Jack W. Pattee, Weihua Guan, James S Pankow, Nathan Pankratz, Weihong Tang – ARIC
  a. The Atherosclerosis Risk in Communities (ARIC) Study has been funded in whole or in part with Federal funds from the National Heart, Lung, and Blood Institute, National Institutes of Health, Department of Health and Human Services, under Contract nos. (HHSN268201700001I, HHSN268201700002I, HHSN268201700003I, HHSN268201700004I, HHSN268201700005I). The authors thank the staff and participants of the ARIC study for their important contributions. Funding was also supported by R01HL103695, R01HL155209, R01HL087641, R01HL059367 and R01HL086694; National Human Genome Research Institute contract U01HG004402; and National Institutes of Health contract HHSN268200625226C. Infrastructure was partly supported by Grant Number UL1RR025005, a component of the National Institutes of Health and NIH Roadmap for Medical Research. Jack W. Pattee was supported by NIH T32GM108557.
15. Michael Levin – PMBB
  a. MGL is supported by the Institute for Translational Medicine and Therapeutics of the Perelman School of Medicine at the University of Pennsylvania and the NIH/NHLBI National Research Service Award postdoctoral fellowship (T32HL007843).
16. Stephen Burgess
  a. SB is supported by a Sir Henry Dale Fellowship jointly funded by the Wellcome Trust and the Royal Society (204623/Z/16/Z). This research was funded by United Kingdom Research and Innovation Medical Research Council (MC_UU_00002/7) and supported by the National Institute for Health Research Cambridge Biomedical Research Centre (BRC-1215-20014). The views expressed are those of the authors and not necessarily those of the National Institute for Health Research or the Department of Health and Social Care.
17. Santhi K Ganesh
  a. SKG is supported by NIH grant R35HL161016, Department of Defense, and the University of Michigan A. Alfred Taubman Institute.
18. Y Eugene Chen
  a. YEC is supported by NIH grant R01-HL109946

