## Supplementary Materials for "Multi-ancestry GWAS deciphers genetic architecture of abdominal aortic aneurysm and highlights *PCSK9* as a therapeutic target"

### Supplementary figures

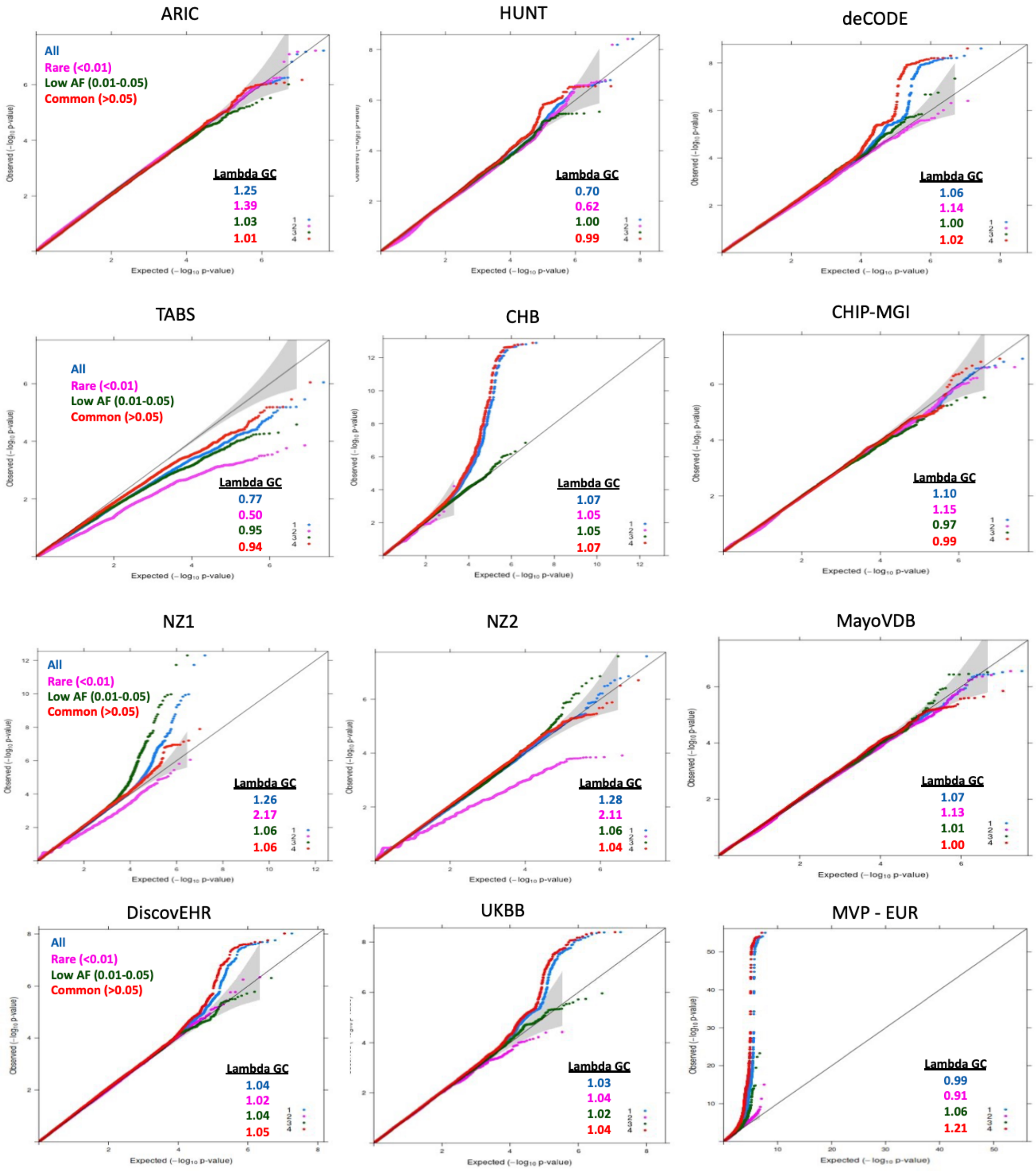

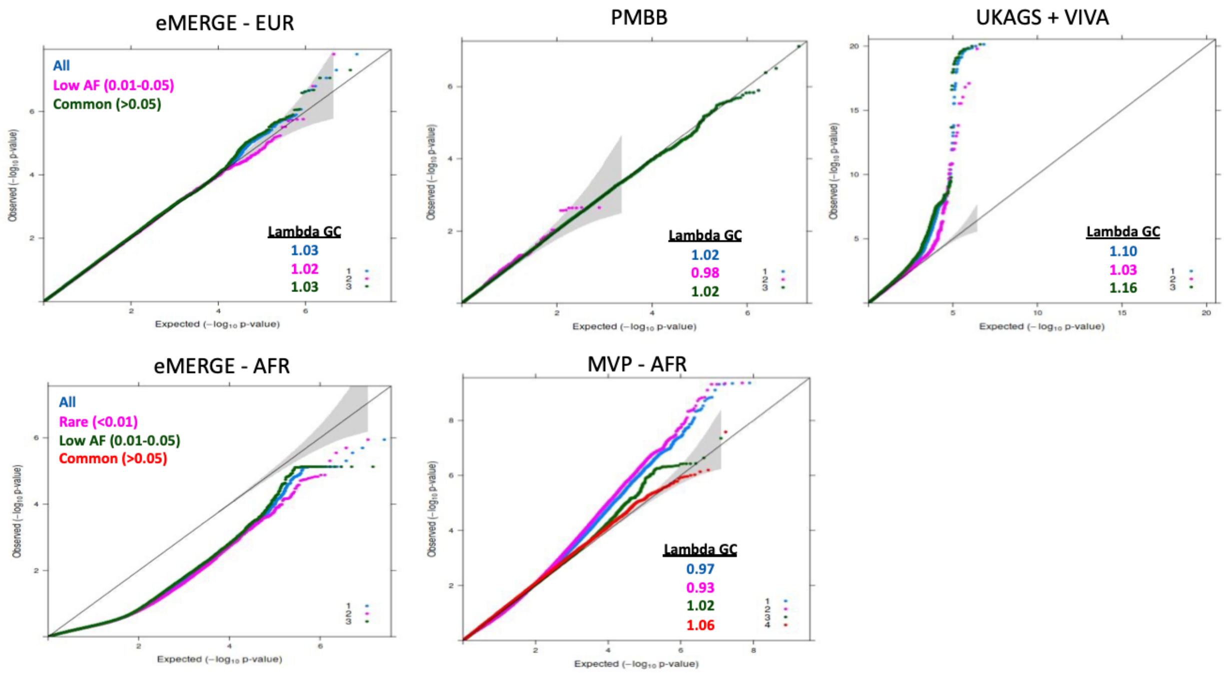


**Figure S1:** QQ plots of AAA GWAS in discovery cohorts.

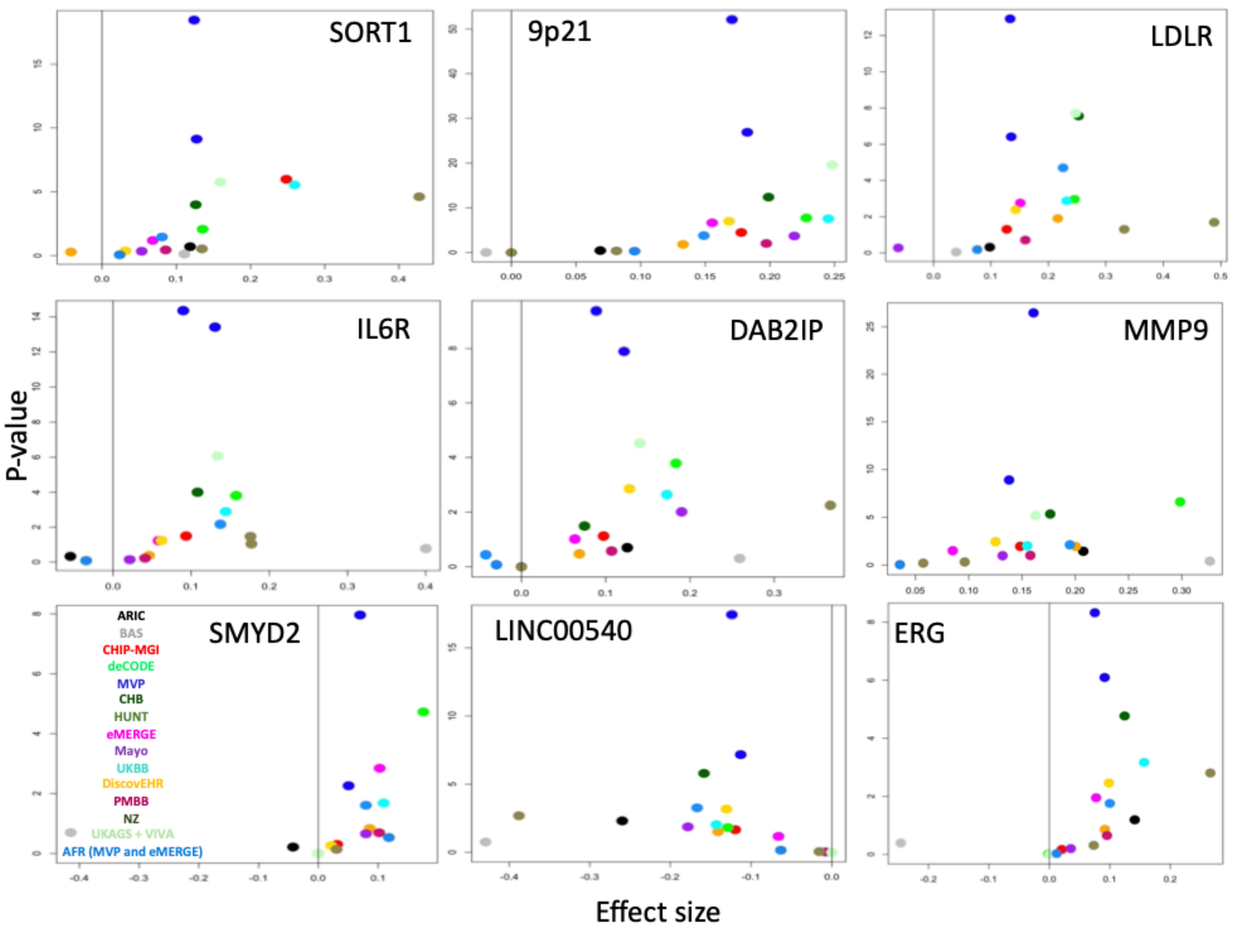


**Figure S2:** Plot of effect size estimation against p-value for 9 known index variants of AAA in discovery cohorts.

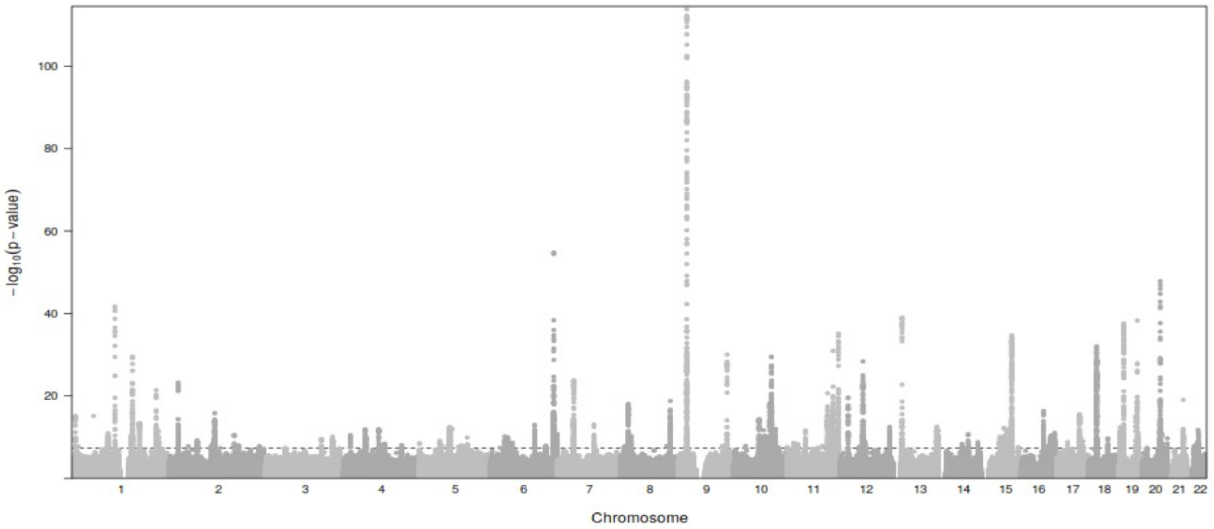


**Figure S3:** Manhattan plot of AAA GWAS meta-analysis.

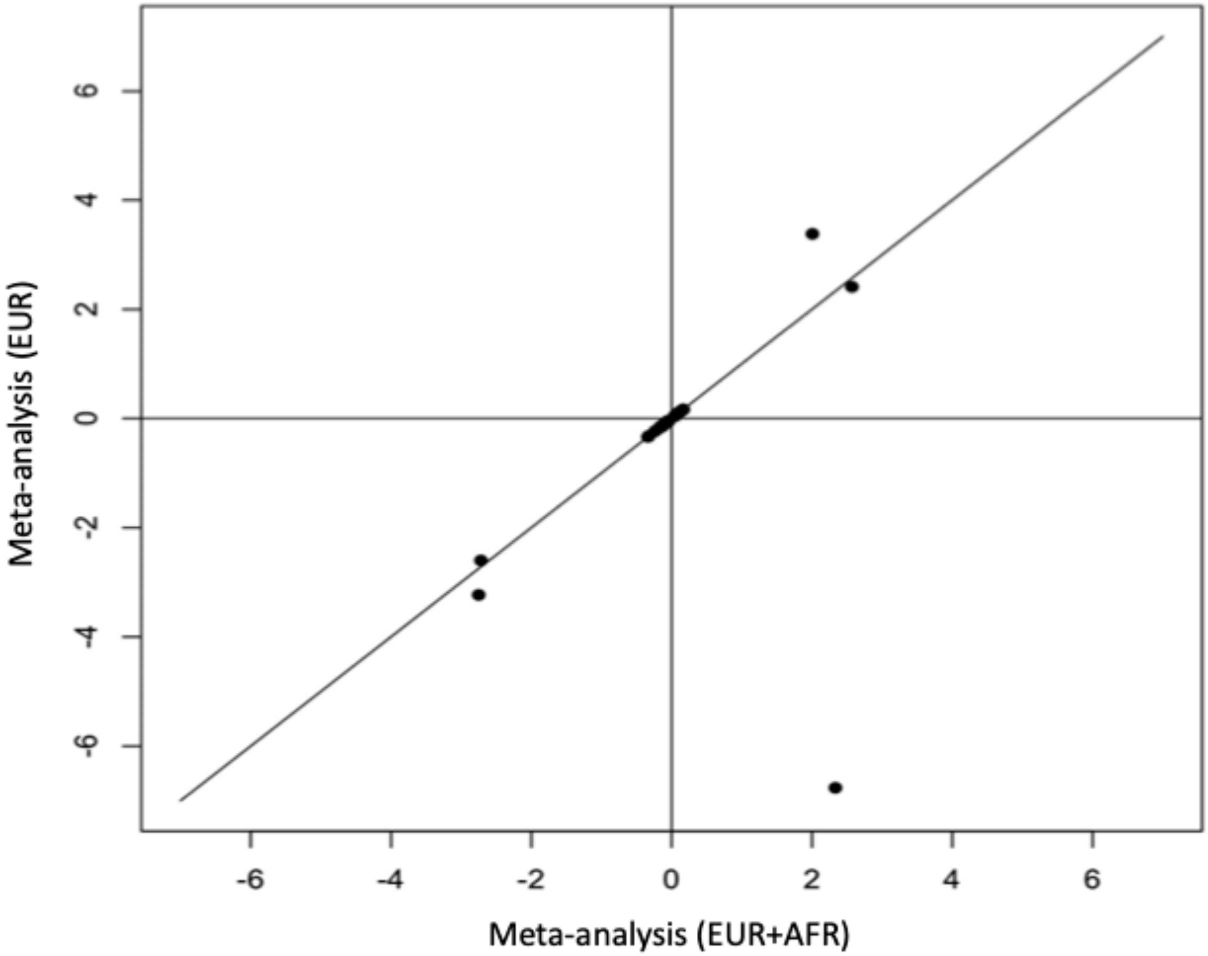


**Figure S4:** Comparison of index variant effect estimates (beta) between meta-analysis with or without AFR ancestry summary statistics. Consistent effect estimates were observed for index variant with MAF > 0.01. Five rare index variants (off-diagonal) were excluded from the follow-up analyses after internal replication step.

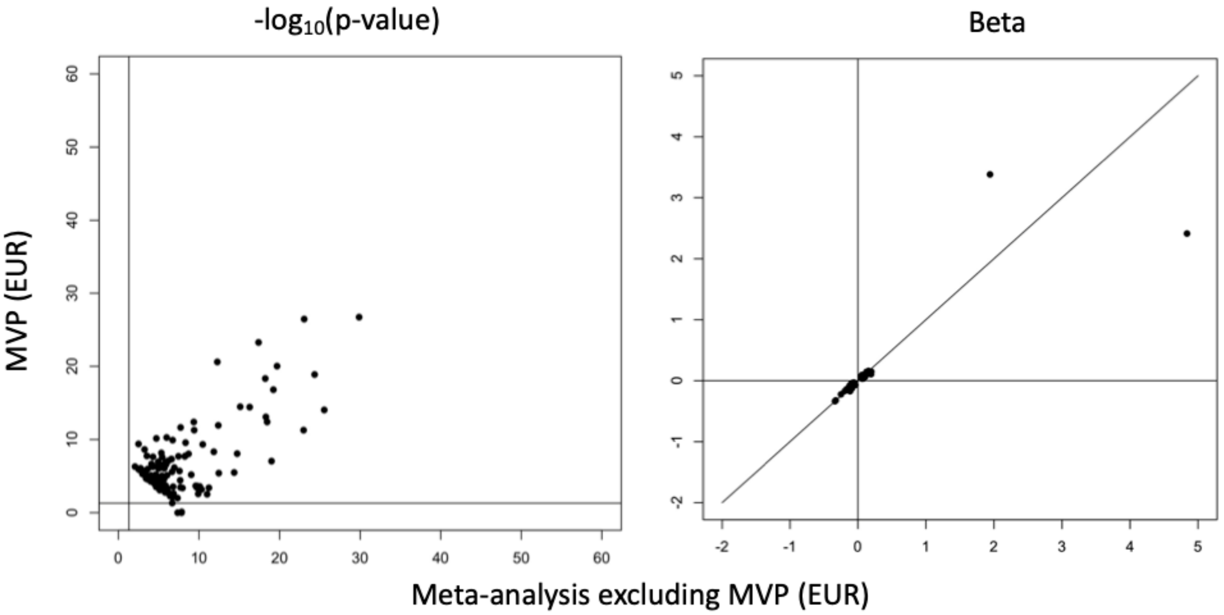


**Figure S5:** Comparison of p-value in 126 genome-wide significant index variants between MVP (EUR) and meta-analysis excluding MVP (EUR). Three index variants have p-value > 0.05 in MVP (EUR) study. Comparison of effect estimate (beta) in the rest of the 123 index variants.

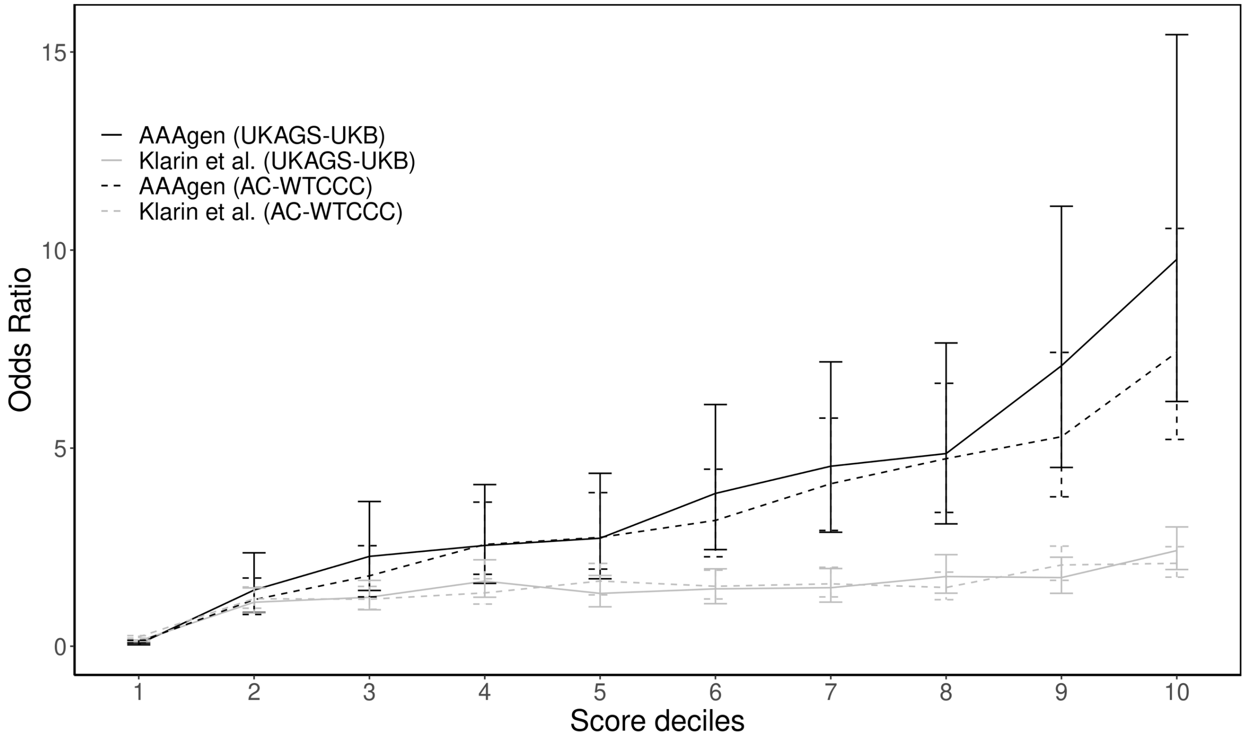


**Figure S6:** Performance of PRS constructed by this meta-analysis (AAAgen) was compared with Klarin et al. (MVP) (Klarin et al., 2020), the largest GWAS of AAA previously published. We observed higher odds ratio by AAAgen compared to Klarin et al. (MVP) in both validation datasets.

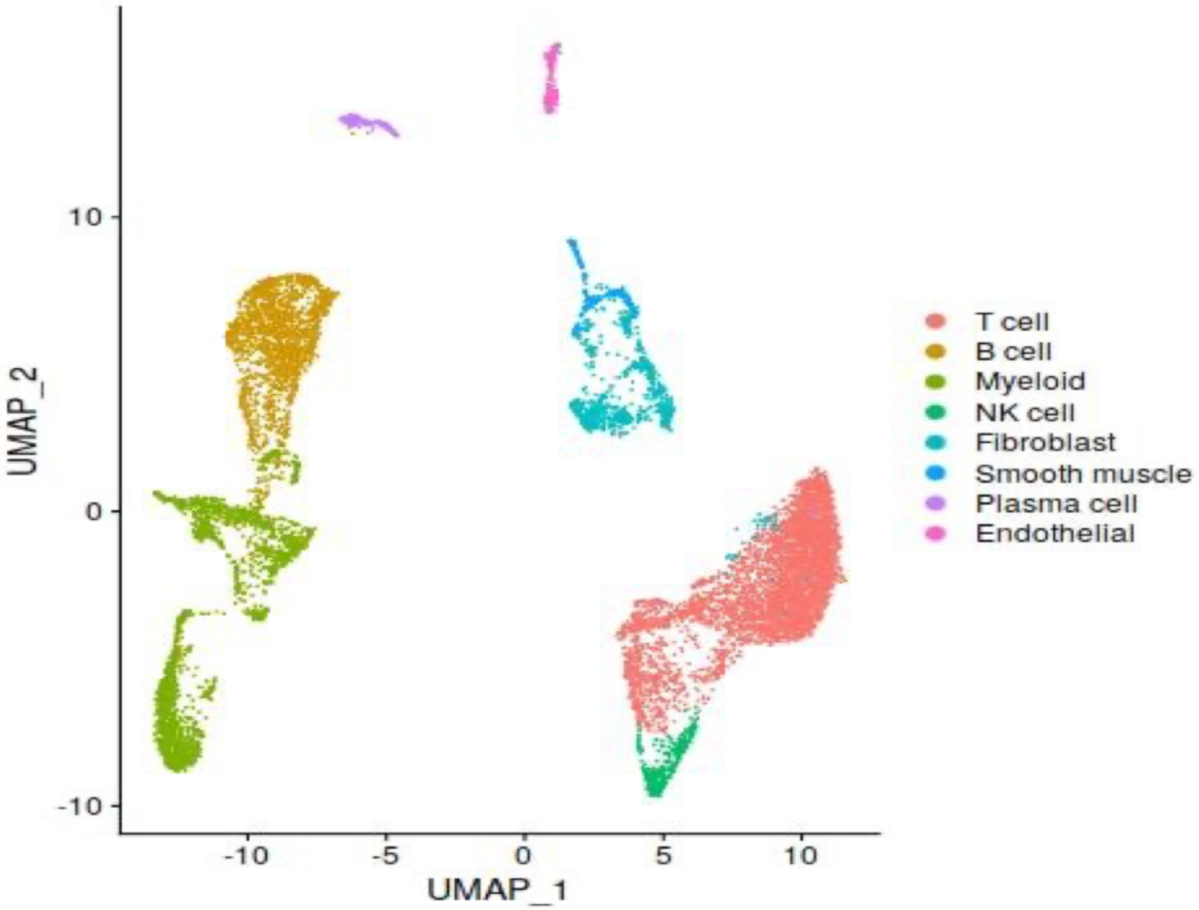


**Figure S7:** UMAP visualization of single cell RNA-seq of aorta used for cell-type enrichment analysis.

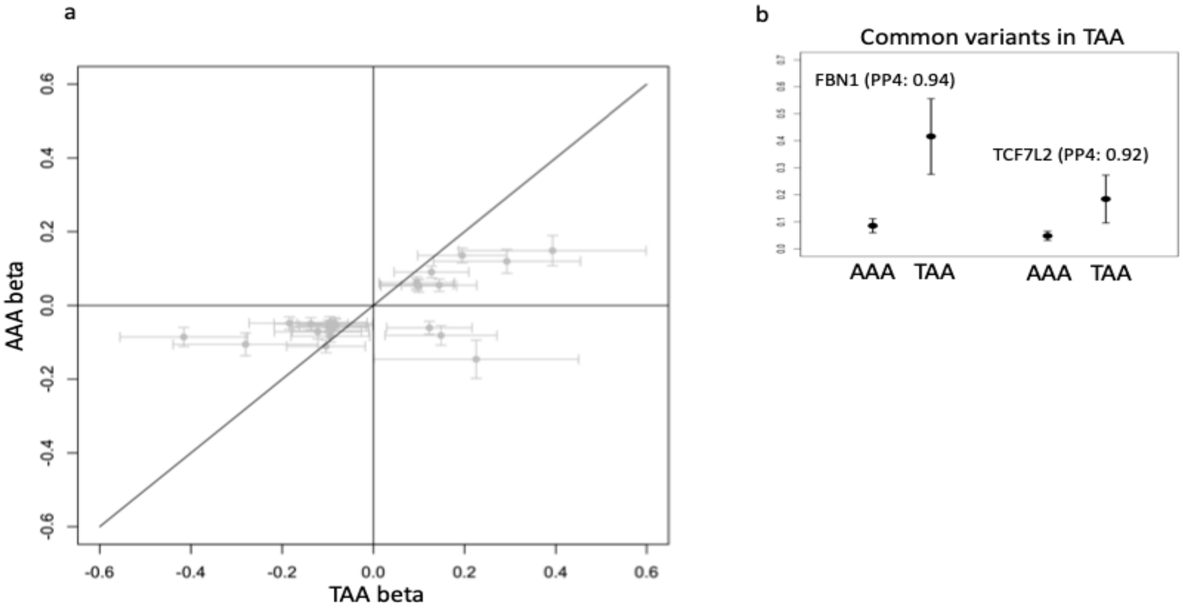


**Figure S8: a)** Comparison of effect estimates (beta) in 24 AAAgen index variants that were observed to have p-value < 0.05 in a recent TAA GWAS (Roychowdhury et al., 2021). **b)** Comparison of effect estimates for two AAA index variants with genome-wide significant TAA effect estimates. PP4 is the posterior probability of the same causal variant in two traits by colocalization.

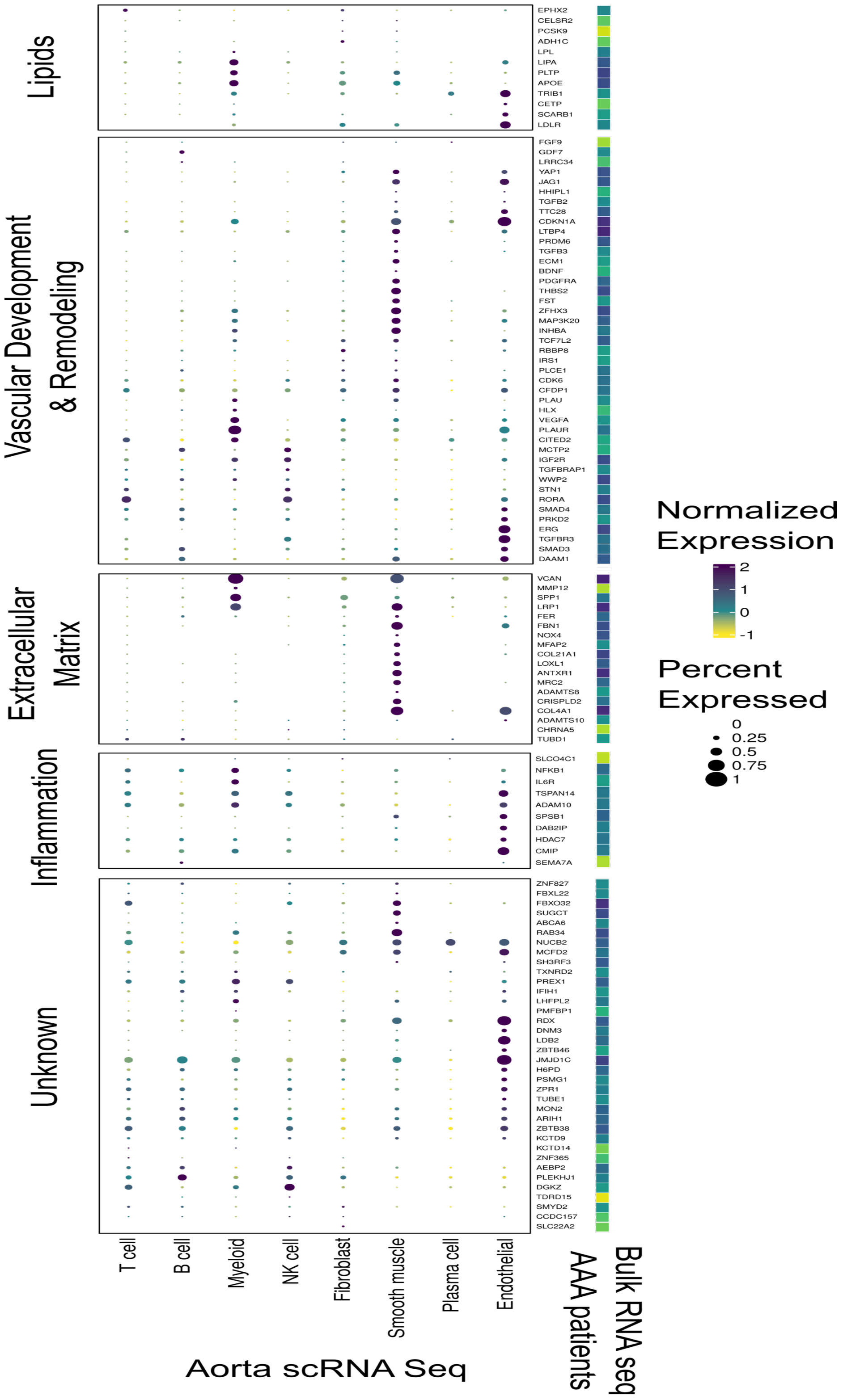


**Figure S9:** Expression of prioritized genes in bulk RNA-seq of 15 AAA patients (right column) and in scRNA seq of aorta (dots in the matrix).

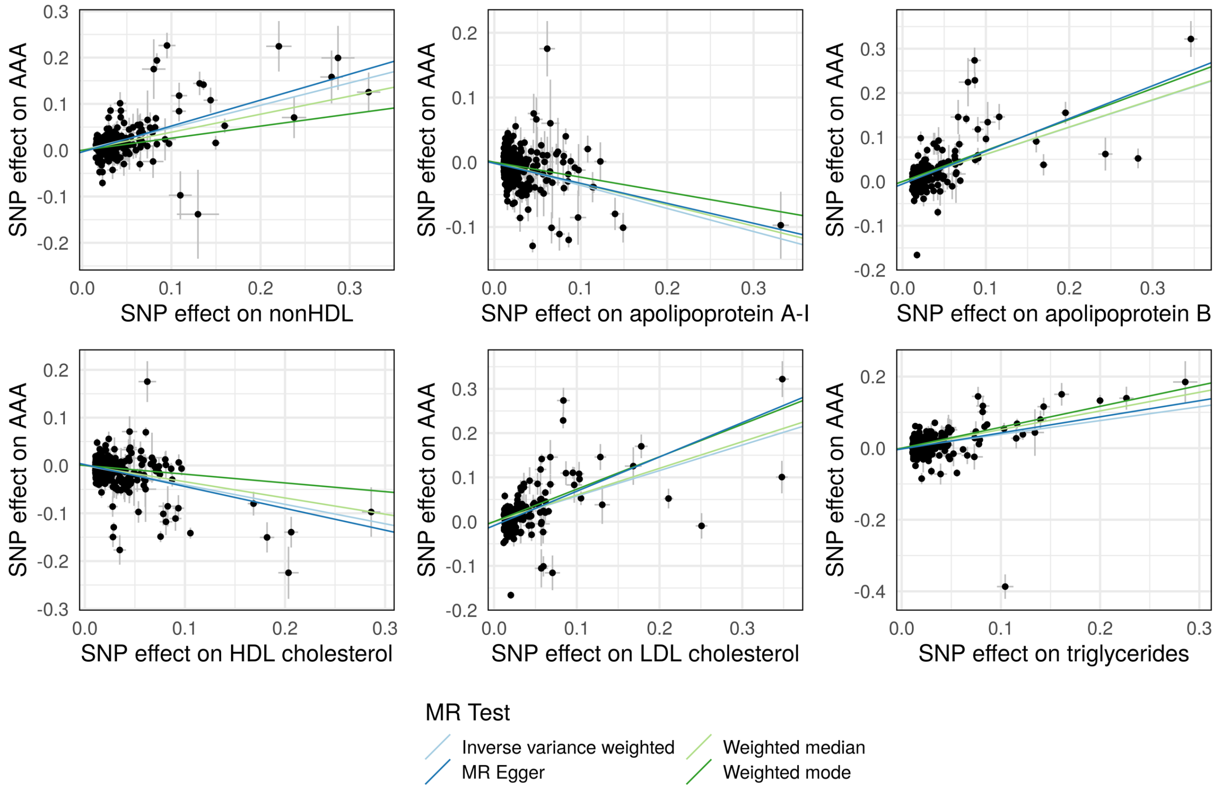


**Figure S10:** Genetic instruments for each lipoprotein-related trait were obtained from GWAS performed with UK Biobank participants. Each point represents the effect of each SNP on AAA (y-axis) and each lipoprotein trait (x-axis). Bars represent standard errors. The slope of each colored line represents the estimated effect of each lipoprotein trait on AAA using MR models which make varying assumptions about the presence of pleiotropy and invalid genetic instruments.

| Residual LDL-c Quintile | OR | 95% CI | P |
|---|---|---|---|
| Quintile 1 (68 mg/dL) | 0.43 | [0.182, 1.01] | 0.05 |
| Quintile 2 (95 mg/dL) | 0.38 | [0.136, 1.07] | 0.07 |
| Quintile 3 (110 mg/dL) | 0.26 | [0.0788, 0.876] | 0.03 |
| Quintile 4 (130 mg/dL) | 0.15 | [0.0368, 0.579] | 0.006 |
| Quintile 5 (170 mg/dL) | 0.25 | [0.0633, 1.02] | 0.05 |

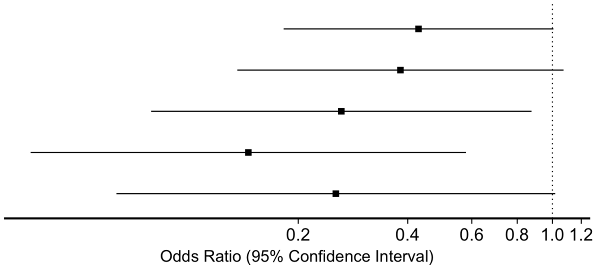


**Figure S11:** Non-linear Mendelian randomization was performed to evaluate the association between LDL-cholesterol lowering and risk of AAA. Across the spectrum of baseline LDL-cholesterol levels, genetically proxied LDL-C-lowering was associated with lower risk of AAA, with no evidence in support of a non-linear relationship.

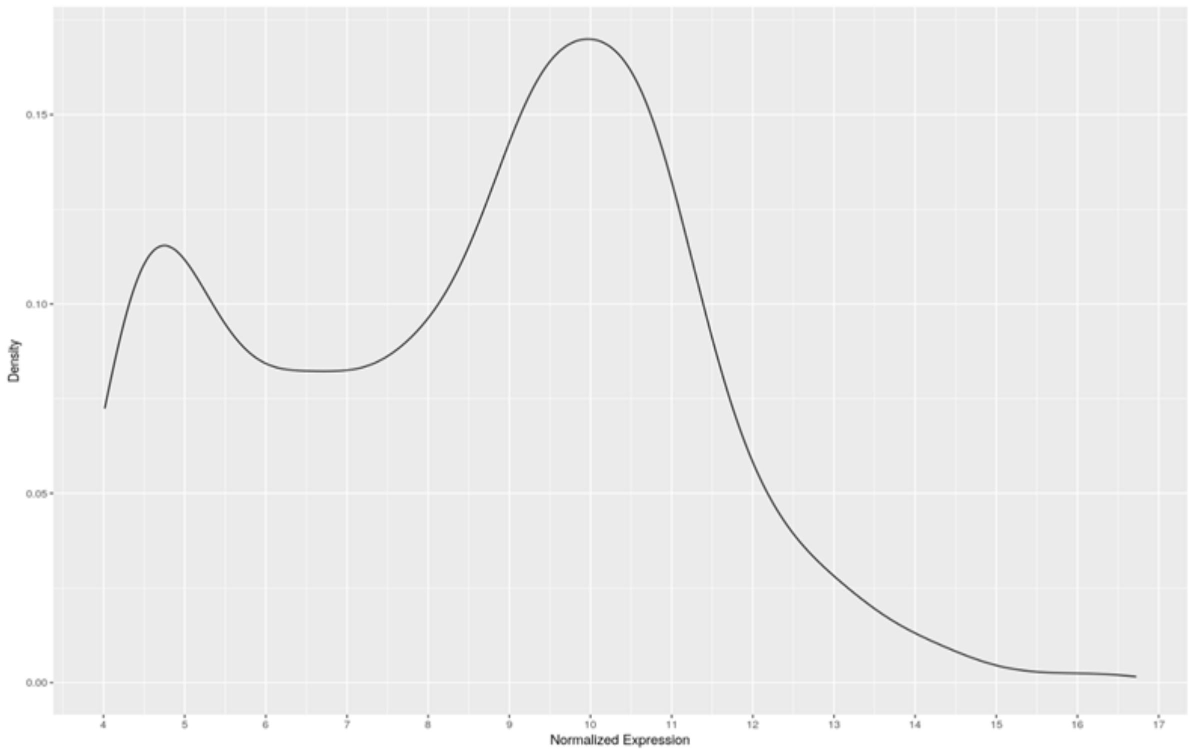


**Figure S12:** Presence or absence of aortic gene expression from bulk RNA-seq of 15 AAA patients was decided based on this bimodal distribution of VST normalized expression (see methods).

### Supplementary Methods

#### Additional details about discovery cohorts

**ARIC:** The Atherosclerosis Risk in Communities (ARIC) Study, a population-based cohort designed to investigate cardiovascular disease and its risk factors, recruited 15,792 individuals aged 45 to 64 years in 1987 through 1989, by list or area probability sampling, from four US communities (Forsyth County, North Carolina; Jackson, Mississippi (African Americans only); the northwest suburbs of Minneapolis, Minnesota; and Washington County, Maryland) (The Atherosclerosis Risk in Communities (ARIC) Study: design and objectives. The ARIC investigators, 1989). ARIC performed a baseline examination among participants in 1987-1989 and followed them by annual telephone contact and multiple reexaminations. All participating institutions gave institutional review board approval for this study and all participants provided written informed consent. ARIC conducted annual telephone calls with participants to ask about any interim hospitalizations and identified deaths, and these records were obtained. ARIC also conducted surveillance of local hospitals to identify additional hospitalizations or deaths. Linkage of participant identifiers with Medicare data from the Centers for Medicare and Medicaid Services (CMS) for 1991-2011 was conducted to find additional hospital or outpatient events for those over 65 years. ARIC identified incident, clinical AAAs by searching hospitalization and death records as well as Medicare data through 2011. Clinical AAA were defined as those who had a hospital discharge diagnosis from any of the above sources, or two Medicare outpatient claims that occurred at least one week apart, with *ICD-9-CM* codes of 441.3 or 441.4, or procedure codes of 38.44 or 39.71, or the following cause of death codes: *ICD-9* 441.3 or 441.4 or *ICD-10* code I71.3 or I71.4 (Folsom et al., 2015; Tang et al., 2016). AAAs based on procedure codes were required to be verified by diagnosis codes. Thoracic, thoracoabdominal, or unspecified aortic aneurysms were treated as non-events. Participants reporting prior AAA surgery or aortic angioplasty at baseline were excluded.

Genomic DNA from blood was genotyped at the Broad Institute with the Affymetrix Genome-Wide Human SNP array 6.0. Genotypes were called using Birdseed software. Participants were excluded if they had a call rate <95% or if their genotype was discordant with known sex or finger-printing genotyping. ARIC conducted race-specific imputation of variant dosages to the 1000 Genomes Project Phase I version 3 reference panel. Before imputation, individuals were removed for being first-degree relatives, genetic outliers, or not matching existing genotype data. The following exclusions were applied to identify a final set of 682,749 autosomal SNPs used for imputation: call rate < 95%, HWE P < $10^{-5}$, MAF < 0.5%. Imputation was performed in two steps: (1) pre-phasing with ShapeIt (v1.r532 ), followed by (2) imputation with IMPUTE2 (Howie et al., 2011; Howie et al., 2009) Phasing with ShapeIt that was run with the parameters: --states-phase 200. Final imputations using IMPUTE2 included the reference panel:1,000 Genomes haplotypes -- Phase I integrated variant set release (v3) in NCBI build 37 (hg19) in chunks of size 5 Mb. All 1092 individuals were used for the imputation from the reference panel. Principal components (PCs) based on the GWAS data were generated by EIGENSTRAT (Price et al., 2006) to reflect the population structure or genetic ancestry of the ARIC participants. Of 11,447 European Americans who were at risk of AAA at baseline in 1987-1989, 8,962 EAs had the GWAS array genotyping and imputation data. Among them, 408 individuals were diagnosed with AAA from 1987-1989 through 2011. Association analysis was performed using logistic regression in SNPTEST (Marchini and Howie, 2010) among 408 AAA cases and 8554 non-AAAs of EUR ancestry. Covariate adjustment included age at baseline, sex, pack-years of smoking, and PCs 1-5.

**CHB-CVDC and DBDS:** Copenhagen Hospital Biobank (CHB) is a hospital driven biobank and includes leftover EDTA blood samples drawn for blood type testing or red cell antibody screening from hospitalized patients in the Danish Capital Region (Sorensen et al., 2021). In addition to genetics, EHR, national socioeconomic and health registries extensively characterize each patient. Patients with AAA are included under the Copenhagen Hospital Biobank Cardiovascular Study (CHB-CVDC) [https://bmjopen.bmj.com/content/11/12/e049709]. The healthy blood donors from "The Danish Blood Donor Study" (DBDS) are included in the study as controls (Hansen et al., 2019). Both CHB-CVDC and DBDS have been approved by the National Committee on Health Research Ethics (NVK-1708829 and NVK-1700407) and the Danish Capital Region Data Protection Office (P-2019-93 and P-2019-99). AAA cases were identified using the following ICD codes (ICD8: 44120/44121/44129; ICD10: I71.3/I71.4). Individuals with aorta dissection were not included in the analysis. AAA cases were compared to the remaining individuals from the DBDS and CHB-CVDC studies excluding individuals with abdominal aortic aneurysm, thoracic aortic aneurysm and intracranial aneurysm using the following ICD codes (ICD8: 44109/44111/44110/44119/44120/44121/44129/44299/43000/43001/43008/43009/43090/43091/43098/43099/43701/43791; ICD10: I71.0/I71.1/I71.2/I71.3/I71.4/I71.5/I71.6/I71.8/I71.9/I72/I60/I67.1). The Infinium Global Screening Array from Illumina was used for genotyping samples from CHB-CVDC and DBDS. Whole-genome sequence data from 8429 Danes along with 7146 samples from North-Western Europe forms a reference panel backbone used for imputation (Helgadottir et al., 2020). The association analysis of 3,079 AAA cases and 180,236 controls was performed with SAIGE using year of birth, sex and 10 PCs as covariates.

**CHIP+MGI:** The Cardiovascular Health Improvement Project (CHIP) is a cohort of individuals treated at Michigan Medicine with linked genotype, EHR, family history data. The Michigan Genomics Initiative (MGI) is a hospital-based cohort with linked genotype and EHR data from participants recruited during pre-surgical encounters at Michigan Medicine. Both studies were approved by the Institutional Review Board of the University of Michigan Medical School (IRBMED) (HUM00052866, HUM00071298) and informed consent was obtained from study participants.

Samples in CHIP and MGI were genotyped using two versions of customized Illumina Infinium CoreExome-24 bead arrays: UM_HUNT_Biobank_11788091_A1 and UM_HUNT_Biobank_v1-1_20006200_A (see Fritsche et al. (Fritsche et al., 2018) for more details). Sample and variant level QC (including joint QC of CHIP and MGI) was performed as in Roychowdhury et al. (Roychowdhury *et al.*, 2021). After QC, 385,816 polymorphic variants from 42,119 samples (CHIP+MGI) were used for imputation using the Michigan Imputation server. Imputation was performed with the HRC panel using Minimac4 (Das et al., 2016). Cases were identified from both CHIP and MGI while controls were identified from MGI. 534 cases from CHIP were identified as aneurysm in abdominal aorta following diagnosis by Cardiologists and after excluding cases with known dissection. From MGI, 749 cases were identified using ICD codes (ICD9: 441.3/4441.4; ICD10: I71.3/I71.4) after excluding cases with known dissection (ICD9: 441.00-03; ICD10: I71.00-03). After removing samples with related phenotypes (phecodes 440–449.99), a case-control matching strategy was used to identify 12,202 controls from MGI. Case-control matching was performed using MatchIt package (Ho et al., 2011) in R using birth year, gender, array version and 4 genotype PCs. Association analysis was performed using SAIGE (Zhou et al., 2018) with birth year, gender, array version and 4 genotype PCs as covariates.

**deCODE:** Icelandic individuals with AAA were identified from a registry of individuals, using ICD codes (ICD9: 441.3, 441.4, ICD10: I71.3, I71.4) who were admitted at Landspitali University Hospital, in Reykjavik, Iceland, 1980–2016, or diagnosed at private clinic in Reykjavik, Iceland. In total, whole genome data from 1,656 subjects with AAA, enrolled as part of the cardiovascular disease (CVD) genetics program at deCODE, were included in this study. The Icelandic controls used (n=265,410) were selected from individuals who have participated in various GWA studies and who were recruited as part of genetic programs at deCODE. Individuals with known cardiovascular disease were excluded as controls but controls were unscreened for AAA.

The preparation of samples and the whole-genome sequencing of 49,962 Icelanders has been described in detail elsewhere (Gudbjartsson et al., 2015a; Jonsson et al., 2017). In short, 37.6 million high-quality sequence variants were identified by sequencing 49,962 Icelanders using GAIIx, HiSeq, HiSeqX, and NovaSeq Illumina technology to a mean depth of at least 17.8×. SNPs and indels were identified and their genotypes called using joint calling with Graphtyper (Eggertsson et al., 2017). Additionally, over 165,000 Icelanders (including all sequenced Icelanders) have been genotyped using various Illumina SNP chips and phased using long-range phasing (Kong et al., 2008), which allows for improving genotype calls using the information about haplotype sharing. The genotypes of the high-quality sequence variants were imputed into the chip-typed Icelanders (Gudbjartsson et al., 2015b). To increase the sample size and power to detect associations, the sequence variants were also imputed into relatives of the chip-typed using genealogic information. All the tested variants had imputation information over 0.8. To test for association of imputed genotypes with AAA we used software developed at deCODE genetics (Gudbjartsson *et al.*, 2015a). In the association analysis, we adjusted for gender, county of origin, current age or age at death (first and second order term included), blood sample availability for the individual, and an indicator function for the overlap of the lifetime of the individual with the time span of phenotype collection. We used LD score regression to account for distribution inflation due to cryptic relatedness and population stratification (Bulik-Sullivan et al., 2015).

**DiscovEHR:** The DiscovEHR cohort is a collaborative effort between Geisinger Health System and the Regeneron Genetics Center. Participants are from Geisinger Health System's MyCode Community Health Initiative including patients from rural Pennsylvania (USA) recruited from 2007-2021. The GHS MyCode initiative and the DiscovEHR study were approved by the Geisinger Institutional Review Board (2006-0258). We included individuals with EHR data, genotype data, and those determined to be of primarily European descent based on SNP-derived principal components analysis.

The Regeneron Genetics Center genotyped DiscovEHR participants on either the Illumina Omni Express Exome or Global Screening Array. Within each array subset, we performed quality control and imputation into the Haplotype Reference Consortium reference panel using the Michigan Imputation Server. We filtered each array subset for MAC>5, site and individual level missingness >10%, HWE p-value $< 1\times10^{-15}$. Following imputation, we merged the two datasets, and removed variants with MAF<0.005 and imputation INFO<0.3. We defined AAA cases based on the presence of at least two instances of any of the following ICD10 codes: 441.3, 441.4, I71.3, I71.4, and required controls to lack any occurrence of the aforementioned ICD10 codes, in addition to the ICD10 codes I71-75, I77-79, K55. We further removed individuals who contributed to the study via the eMERGE consortium. Final analysis included 2,238 cases and 105,433 controls. We performed association analysis using whole genome regression in REGENIE (v1.0), including Age, $Age^2$, Sex, Age×Sex, $Age^2$×Sex, 10 common variant (MAF>1%) derived principal components, and an array batch indicator as covariates in the model.

**eMERGE:** The electronic Medical Records and Genomics (eMERGE) network is a consortium that consists of EHR data on approx. 100,000 patients from twelve institutions across the US linked with their genetic data. eMERGE consists of samples from diverse ancestral background with approx. 20% samples from African ancestry. In the eMERGE network (Phase 3 ascertainment), this phase of the eMERGE Network was initiated and funded by the NHGRI through the following grants: U01HG8657 (Kaiser Washington/University of Washington); U01HG8685 (Brigham and Women's Hospital); U01HG8672 (Vanderbilt University Medical Center); U01HG8666 (Cincinnati Children's Hospital Medical Center); U01HG6379

(Mayo Clinic); U01HG8679 (Geisinger Clinic); U01HG8680 (Columbia University Health Sciences); U01HG8684 (Children's Hospital of Philadelphia); U01HG8673 (Northwestern University); U01HG8701 (Vanderbilt University Medical Center serving as the Coordinating Center); U01HG8676 (Partners Healthcare/Broad Institute); and U01HG8664 (Baylor College of Medicine). In eMERGE network (Phase 1 and 2 ascertainment), the eMERGE Network was initiated and funded by NHGRI through the following grants: U01HG006828 (Cincinnati Children's Hospital Medical Center/Boston Children's Hospital); U01HG006830 (Children's Hospital of Philadelphia); U01HG006389 (Essentia Institute of Rural Health, Marshfield Clinic Research Foundation and Pennsylvania State University); U01HG006382 (Geisinger Clinic); U01HG006375 (Group Health Cooperative/University of Washington); U01HG006379 (Mayo Clinic); U01HG006380 (Icahn School of Medicine at Mount Sinai); U01HG006388 (Northwestern University); U01HG006378 (Vanderbilt University Medical Center); and U01HG006385 (Vanderbilt University Medical Center serving as the Coordinating Center) with U01HG004438 (CIDR) and U01HG004424 (the Broad Institute) serving as Genotyping Centers.

Samples in the eMERGE Phase III were genotyped on 78 different Illumina and Affymetrix SNP array platforms. Data from different sites and genotyping platforms were imputed using Haplotype Reference Consortium reference panel on Michigan Imputation Server and then merged into one set (Stanaway et al., 2019; Verma et al., 2014). Samples with 2 or more instances of ICD9 codes 441.3/4441.4 or ICD10: I71.3/I71.4 were defined as cases and samples without instance of case ICD codes were defined as controls. Analysis was separated by samples of European (EUR) and African (AFR) ancestry. Case control matching was performed on EUR and AFR samples to retain 5 controls per each case, matching based on age and sex using MatchIt R program. We identified 3092 cases and 15,025 controls in the EUR dataset and 119 cases and 603 controls in the AFR dataset. SNPs with imputation R2>0.3, SNP call rate >99% and MAF >1% were retained for the analyses, samples with sample call rate less than 99% were removed. Logistic Regression on unrelated samples was performed using PLINK 2.0, all models were adjusted by clinical site and first 5 principal components.

**HUNT**: The Trøndelag Health (HUNT) Study (Krokstad et al., 2013) is a population-based health study from the Trøndelag region in Norway. Individuals have been enrolled in the study though multiple phases (HUNT1, HUNT2, HUNT3, and HUNT4) beginning in 1984. During each enrollment period, individuals living in the region who are 20 years or older are invited to participate. To date, over 120,000 individuals have participated and have longitudinal health record data available from surveys, medical records, and biological samples.

Approximately 70,000 HUNT participants have been genotyped using the Illumina HumanCoreExome array. Following standard QC, the samples were phased with Eagle2 v2.3 (Loh et al., 2016b) imputed with Minimac3 (Das *et al.*, 2016) using a combined reference panel consisting of the Haplotype Reference Consortium v1.1 combined with whole genome sequencing of 2,201 HUNT participants. AAA cases were defined by ICD-9 codes 441.3 and 441.4 and ICD-10 codes I71.3 and I71.4. A total of 734 cases and 68,901 controls were included in the analysis. Association testing was performed using SAIGE (Zhou *et al.*, 2018) with batch, sex, birth year, and PC1-4 as covariates.

**Mayo VDB:** Mayo Vascular Disease Biorepository (VDB) at the Mayo Clinic was established to archive DNA, plasma, and serum from patients with suspected atherosclerotic cardiovascular disease (ASCVD) phenotypes including coronary artery disease, carotid artery stenosis, cerebrovascular disease, peripheral artery disease, abdominal aortic aneurysm, and other vascular conditions (Ye et al., 2013). Mayo VDB enrolled patients referred for noninvasive vascular evaluation and exercise stress testing at the Mayo Clinic Gonda Vascular Center from January 14, 2006 to July 24, 2020. ASCVD phenotypes and related comorbidities were ascertained using previously validated electronic phenotyping algorithms based on both structured data elements (ICD diagnosis codes, CPT codes, and laboratory measurements) and natural language processing of unstructured data elements, such as vascular laboratory and imaging reports in the EHR. The study was approved by the Mayo Clinic Institutional 74 Review Boards (IRB # 08–008355).

Genotyping was performed using three different Illumina platforms, including Illumina Human660W-Quad V1, HumanCoreExome Beadchip, and Human 610 Quad V1 platforms. We combined samples of different platforms by genotype imputation based on a human reference consortium (HRC, r1.1) panel, which includes 64,976 human haplotypes at 39,235,137 SNPs constructed using whole-genome sequencing data from 20 studies of predominantly European ancestry. Before imputation, we excluded rare variants (MAF<0.05 in EUR of 1000 Genome Projects) in the HumanCoreExome and 610 Quad V1 platforms. In addition, a pre-imputation step was conducted, including checking strand, alleles, position, Ref/Alt assignments and frequency differences, as suggested in http://www.well.ox.ac.uk/~wrayner/tools/. For each dataset, we used the Michigan Imputation Server Minimac3 and Eagle2 v2.3 for imputation and phasing from the HRC panel. Following imputation, we combined the individual variant calling format (VCF) file from different datasets for each chromosome. We excluded SNPs with 3+ alleles due to incapability of processing these markers in PLINK format. After combining, we obtained 38,909,200 SNPs (including monomorphic variants) in 9,871 samples in the entire Mayo VDB study. We removed samples that did not pass quality control (n=108) as well as contaminated samples (n=4). Based on identity-by-descent analysis implemented in PLINK, we removed one sample of low genotyping rate from the duplicated samples (n=194) and the first-degree relatives (i.e., parents/children and full siblings, n=204). Using the 1000 Genomes Project, we performed principal component analysis to detect samples of divergent ancestry with the 'smartpca' program in the EIGENSOFT package. Outliers (n=139) that are more than six standard deviations beyond the European samples (i.e., five European populations [CEU, TSI, FIN, GBR, and IBS] in the 1000 Genomes Project) mean score (PC1 and PC2) were excluded. Finally, 10 samples without clinical numbers in the EHR were excluded. In total, we excluded 635 samples in the QC steps. Following genotype data QC, a total of 9,236 individuals are present in the Mayo VDB genotype dataset. After exclusion of overlapping samples between Mayo VDB Dataset and eMERGE III V3 Imputed Array Dataset (Stanaway *et al.*, 2019), AAA cases (n=771) were defined as having an infrarenal abdominal aortic diameter ≥3 cm or a history of open or endovascular AAA repair based on an electronic phenotyping algorithm using Natural Language Processing on ultrasound reports and were also confirmed with manual chart review. Controls (n=4,913) were not known to have AAA and had no ICD-9 diagnosis codes for AAA. These subjects were genotyped on Illumina HumanCoreExome Beadchip, and Human 610 Quad V1 platforms. Association testing was performed on the imputed dataset using SAIGE software with the following covariates: study enrollment age, sex, genotyping platform, first 5 principal components of ancestry.

**MVP:** In the Million Veteran Program (MVP), individuals aged 18 to over 100 years have been recruited from 63 VA Medical Centers across the United States. MVP received ethical/study protocol approval by the VA Central Institutional Review Board, and informed consent was obtained for all participants. Each additional study received approval from their local institutional review board.

DNA extracted from whole blood was genotyped in MVP using a customized Affymetrix Axiom biobank array, the MVP 1.0 Genotyping Array. Veterans (U.S. military personnel) of two mutually exclusive ethnic groups were identified for analysis: 1) non-Hispanic whites (European ancestry), and 2) non-Hispanic blacks (African ancestry) using the HARE algorithm (Fang et al., 2019). Prior to imputation, variants that were poorly called or that deviated from Hardy-Weinberg equilibrium or their expected allele frequency based on reference data from the 1000 Genomes Project (Genomes Project et al., 2015) were excluded. After pre-phasing using SHAPE-IT4 (Delaneau et al., 2019), genotypes from the African Genome Resources reference panel were imputed into MVP participants via Minimac4 software (Howie et al., 2012). Ethnicity-specific principal component analysis was performed using the EIGENSOFT v6 software (Price *et al.*, 2006). In MVP, sample and variant quality control was performed as previously described (Klarin et al., 2018). In brief, duplicate samples, those with more heterozygosity than expected, an excess (>2.5%) of missing genotype calls, or discordance between genetically inferred sex and phenotypic gender were excluded. In addition, one individual from each pair of related individuals (kinship > 0.0884 as measured by the KING 2.0 software) were removed. From the participants passing quality control in MVP, individuals were defined as having AAA or being a disease-free control using a previously utilized (Klarin *et al.*, 2020) definition initially proposed by Denny et al (Denny et al., 2013). AAA cases were defined as the presence of two instances of any of the following ICD-9/10 codes in a participant's EHR: 441.3, 441.4, I71.3, I71.4. Controls were defined as possessing zero occurrences of the aforementioned ICD codes, as well as zero occurrences of the ICD-9 codes 440-448, or ICD-10 codes I71-75, I77-79, K55. In our MVP analysis, we evaluated 17,672 AAA cases and 303,695 controls of European ancestry, and 1,888 AAA cases and 87,728 controls of African ancestry. Genotyped and imputed DNA sequence variants in individuals were tested for association with AAA using logistic regression adjusting for age, sex, and 5 principal components of ancestry assuming an additive model using the PLINK2.0 statistical software program.

**NZ AAA Genetics Study:** The Vascular Research Consortium of New Zealand recruited New Zealand men and women with a proven history of AAA (infra-renal aortic diameter ≥ 30 mm proven on ultrasound or CT scan). Approximately 80% had undergone surgical AAA repair (typically AAA's > 50-55 mm in diameter). The vast majority of cases (>97%) were of Anglo-European ancestry. The control group underwent an abdominal ultrasound scan to exclude (>25 mm) concurrent AAA and Anglo-European ancestry was required for inclusion. Controls were also screened for peripheral artery disease (PAD; using ankle brachial index), carotid artery disease (ultrasound) and other cardiovascular risk factors. All participants were genotyped in two separate case-control cohorts using the Affymetrix SNP6 (cohort 1; 608 AAA cases and 612 controls) or Illumina Omni2.5 (cohort 2; 397 cases and 384 controls) GeneChip arrays and had call rates >95% (mean 99.2%). Imputation was then conducted using IMPUTE 2.2 run on the BCISNPmax database platform (version 3.5, BCI Platforms, Espoo, Finland). The reference haplotypes were based on the 1000 Genomes June 2011 release. Imputed calls were filtered by quality score (>0.9) to restrict to higher quality imputed SNPs. The genomic inflation factors were 1.06 and 1.05 respectively (MAF >0.05, 5.4 million SNPs). Case-control genome wide association analyses were conducted using PLINK (version 1.07).

**PMBB:** Penn Medicine biobank (PMBB) recruits patients from throughout the University of Pennsylvania Health System for genomic and precision medicine research. Participants actively consent to allow the linkage of biospecimens to their longitudinal EHR. Currently, >60 000 participants are enrolled in the PMBB. A subset of ~45000 individuals who have undergone whole exome sequencing and genotyping, performed through a collaboration with the Regeneron Genetics Center. A further subset of ~23000 subjects with imputed genotype data was used in this analysis. 19,515 recruits were genotyped on three different arrays, Illumnia Quad Omni, Global Screening Assay v1 and Global Screening Assay v2. Genotype imputations were performed using the Eagle2 (Loh et al., 2016a) and Minimac softwares (Das *et al.*, 2016) on the Michigan Imputation server and were completed for all autosomes with the HRC reference panel. AAA cases were found using the AAA phecode definition of 442.11, meaning at least 1 encounter with abdominal aortic aneurysm diagnosis codes and excluding other confirmed disease of the arteries. Controls were defined as all others with genotype data and no evidence of AAA. GWAS of 388 cases and 9879 controls was performed using plink software with age at recruitment, gender, genetic determined ancestry and the first 10 principal components.

**TABS:** The Triple A Barcelona Study (TABS) is a hospital-based study recruiting individuals with AAA treated in the Hospital de la Santa Creu i Sant Pau in Barcelona, Spain. All individuals have repetitive measurements of the abdominal aortic diameter, either by CT-scan or by ultrasound along with anthropometric and clinical information. DNA, RNA, and plasma samples were collected from all individuals. All participants gave written informed consent. All procedures were approved by the Institutional Review Board at Hospital de la Santa Creu i Sant Pau.

Genome-wide genotyping was performed using the Infinium Global Screening Array-24 v2.0 from Illumina (coverage 665,608 variants). The genotyping service was carried out at CEGEN-PRB3-ISCIII, and it is supported by grant PT17/0019, of the PE I+D+i 2013-2016, funded by ISCIII and ERDF. Prior to imputation, we performed several quality control filters as follows: gender mismatches, sample call rate (> 95%), SNP call rate (> 98%), heterozygosity test (median plus/ minus 3 times the interquartile range), and HWE test (p-value > $10^{-6}$). We also filtered out non-European individuals by principal component analysis and removed monomorphic markers. All pre-imputation quality control was performed using PLINK 1.90. All chromosomes were then imputed to the HRC reference panel using the Michigan server. After imputing, variants with imputation quality <0.3 or MAF <0.002 were excluded. Cases were defined as those with dilation of the abdominal aorta with a diameter higher than 30 millimeters. Type B dissections that progress to AAA, saccular aneurysms or thoracic aneurysms were excluded. In addition to 42 cases from TABS, 10 cases and 82 controls were leveraged from the Triple A Genetic Study (TAGA) (Peypoch et al., 2020). Also, 401 controls were leveraged from the RETROVE study (Vazquez-Santiago et al., 2017). Association analysis was performed using SAIGE [30104761] with birth year, sex, batch and PCs as covariates.

**UKAGS+VIVA and UKBB:** The UK Aneurysm Growth Study (UKAGS), The Viborg Vascular (VIVA) and UK Biobank (UKBB) cohorts are described together since cases from each cohort were compared with controls from UKBB. Case cohorts were defined by genotyping batches with cases from part of the UKAGS study and the VIVA trial being genotyped together (the remaining UKAGS and VIVA cases being genotyped as a separate batch that was only available after the discovery analysis and was used for polygenic risk score validation together with other new cohorts only available after discovery analysis, see below) and UKBB cases being genotyped as part of that study. The overall sample of UKBB controls was split to provide independent control data for each case dataset.

UKAGS is a prospective study of men attending the NHS aneurysm screening programmes in the UK. Men were recruited into the study after they had been found to have an abdominal aortic aneurysm (AAA) in one of the UK NHS AAA screening programmes and were consented by post for all study activities by the research group at University of Leicester. Men recruited into the UKAGS completed a postal questionnaire to obtain information on smoking, comorbidities and medications. Screening outcomes (ultrasound measured AAA diameter) were obtained directly from the AAA screening programmes. Ethical approval was granted by an NHS research ethics committee. The study was funded by the British Heart Foundation (CS/14/2/30841 and RG/18/10/33842) and the Circulation Foundation.

VIVA screening trial is a randomized, clinically controlled study designed to evaluate the benefits of vascular screening and modern vascular prophylaxis in a population of 50,000 men aged 65-74 years, randomized to either receive an invitation for vascular screening or being a control. Enrolment started October 2008 with major follow-up at 3, 5 and 10 years. Ethical approval was granted by the research ethics committee of Mid Denmark (M20080028), and funded by the FP7, EU, and the Region of Mid Denmark. ClinicalTrials.gov NCT00662480.

UKBB is a large prospective study with over 500,000 participants aged 40–69 years when recruited in 2006–2010. UKBB is available for open access, without the need for collaboration, to any bona fide researcher who wishes to use it to conduct health-related research for the benefit of the public. An independent Ethics and Governance Council oversees adherence to the Ethics and Governance Framework. To identify AAA cases in UKBB the following ICD and OPCS codes were searched for in the UKBB hospital inpatient data. Any individuals with one or more of the following codes was defined as a case: ICD9: 441.3, 441.4; ICD10: I71.3, I71.4; OPCS4: L184, L185, L186, L194, L195, L196, L271, L275, L276, L281, L285, L286. Controls were selected from UKBB so as to exclude individuals with any potential aortic pathology such as thoracic aortic aneurysm using the following ICD codes as exclusion criteria: ICD9: 441.00, 441.01, 441.02, 441.03, 441.1, 441.2, 441.5, 441.6, 441.7, 441.9; ICD10: I71.0, I71.1, I71.2, I71.5, I71.6, I71.8, I71.9. The group of controls was then restricted to the UKBB 'in white British ancestry subset'. Any individuals whose sex did not match the inferred gender were excluded and one of any pairwise kinships excluded. The derived pool of control samples was then used to select age and sex matched control groups (without ovelap) for each of the separate case groups used in the discovery and validation GWAS (see details below).

All participants in UKAGS and VIVA were genotyped using the UKBB Axiom Array. The following QC filters were used: 1) Males 65 and over, 2) white British/Danish ancestry only, 3) one of any pairwise kinships removed, 4) outliers identified by PCA and excluded, 5) batch missing test (--test-missing in PLINK) to exclude variants with highly significant difference in missingness between cases and controls (P <0.001), 6) variants with missing call rates > 2% excluded. A pre-imputation check against the 1000G reference SNP list was carried out with the McCarthy Group perl tool HRC-1000G-check-bim-v4.3.0 and the 1000GP_Phase3_combined.legend file, which was used to check and correct strand, position and ref/alt assignments and remove A/T & G/C SNPs if MAF > 0.4 and SNPs with differing alleles (https://www.well.ox.ac.uk/~wrayner/tools/). All imputation, including for UKBB was carried out on the Michigan Imputation Server, Minimac 4, 1000G Phase3 v5 (GRCh37/hg19) reference panel (Rsq filter = 0.3). Association analysis of 3595 cases (3209 UKAGS + 386 VIVA) and 15773 controls (UKBB) was performed with PLINK v2.00a, with following parameters, --geno 0.02, --hwe 1e-8, --maf 0.01.

UKBB participants were genotyped using the UKBB Axiom array and UKBB BiLEVE Axiom array. This analysis made use of the available UKBB imputed data, imputed with HRC and UK10K + 1000 Genomes panels. The following QC filters were used: **1)** Age and sex of controls matched to that occurring in cases (age range: 52-79), 2) white British ancestry only, 3) one of any pairwise kinships removed, 4) samples missing X / XY data excluded, 5) MAF > 0.01, 6) Info score > 0.5. Association analysis of 1241 cases (1081 males, 160 females) and 6276 controls (5466 males, 810 females) was performed with SNPTEST v2.5.2, with the following parameters, -frequentist additive, -method expected.

**Supplementary Methods for Conventional Mendelian Randomization**

We performed conventional two-sample Mendelian randomization to estimate the total effect of each major lipoprotein-related trait on AAA. We constructed genetic instruments from independent ($r^2$ < 0.001, distance > 10,000kb) genetic variants associated with each major lipoprotein-related trait in the UK Biobank. After identifying the corresponding genetic variants in our GWAS of AAA and harmonizing the effect alleles, we performed MR using the

*TwoSampleMR* package in R (Hemani et al., 2018). Our primary analysis used the inverse variance weighted method. In sensitivity analyses, we performed MR-Egger, weighted median, and weighted mode MR, which make different assumptions about the presence of pleiotropy (Davies et al., 2018).

**Supplementary Methods for MR-BMA.**

We performed a variable selection method in a multivariable Mendelian Randomization (MR) framework to prioritize the causal lipoprotein determinants of the outcomes. Multivariable MR extends the basic MR framework to include multiple exposures in one joint model, accounting for horizontal pleiotropy among exposures, which is particularly relevant when considering highly correlated traits like blood lipoprotein-related traits as exposures (Burgess and Thompson, 2015). In order to rank and select the likely causal lipoprotein risk factors for AAA, we employed an extension of multivariable MR called Mendelian randomization Bayesian model averaging (MR-BMA), a Bayesian approach for prioritizing causal exposures in a two-sample multivariable MR setting (Zuber et al., 2020). MR-BMA performs variable selection by evaluating models with all possible combinations of lipoprotein-related traits as exposures and computing the posterior probability that the model contains the true causal risk factors. Unlike other univariate or multivariable MR methods, MR-BMA aims to identify true causal risk factors among correlated traits, rather than estimate the magnitude of effect. The marginal inclusion probability (level of evidential support for each exposure) is derived from the sum of all posterior probabilities of the models where the specific exposure was included. We removed influential variants based on the Cook's distance and outliers based on the q-statistic as previously recommended (Zuber et al., 2021). An empirical permutation procedure was performed to calculate p-values. Briefly, the expected marginal inclusion probability distribution for each risk factor under the null hypothesis was generated by performing 1,000 permutations of the MR-BMA analysis, holding the SNP-risk factor associations constant and randomly permuting the SNP-outcome associations. The observed marginal inclusion probabilities for each risk factor were then compared to the expected distribution under the null, with p-values computed by $p_j = (r_j + 1)/(nperm + 1)$, where rj represents the rank of the observed marginal inclusion probability of a given risk factor (j) across all permutations (nperm = 1000). Adjustment for multiple testing was done using the Nyholt correction for correlated traits.

**Supplementary Methods for Nonlinear Mendelian Randomization**

Typically, single- and two-sample Mendelian randomization analyses assume a linear relationship between the exposure of interest and outcome. Prior meta-analysis has suggested a potential interaction between baseline LDL-cholesterol and PCSK9-inhibition, where individuals with high baseline LDL-c derive outsize cardiovascular benefit from PCSK9-inhibition (Khan et al., 2019). Here, we explored the relationship between genetically proxied LDL-cholesterol lowering by PCSK9-inhibitors and AAA, with a focus on evaluating the possibility of a non-linear treatment effect. We constructed a genetic instrument with uncorrelated ($r^2 < 0.01$, distance = 10,000kb) LDL-cholesterol-associated variants ($p < 5x10^{-8}$) located +/- 250kb from the PCSK9 locus to proxy the effects of PCSK9-inhibition (Levin et al., 2021). Among European-ancestry participants of the VA Million Veteran Program with available genotype, phenotype, and LDL-cholesterol lab values, we performed non-linear Mendelian randomization (Burgess et al., 2014; Sun et al., 2019). Individuals were first stratified into quantiles by residual LDL-cholesterol levels after accounting for the effects of the genetic variants included in the PCSK9 genetic instrument. Within each quintile, local average causal effects were estimated using the ratio of effects MR method. Linearity of the resultant local average causal effects was assessed using the heterogeneity and fractional polynomial methods implemented in *nlmr* package in R (Staley and Burgess, 2017), with $p < 0.05$ set as the threshold for significance.

**Nonlinear Mendelian Randomization for *PCSK9***

We additionally evaluated whether there was a linear effect observed for LDL cholesterol reduction via the *PCSK9* gene pathway. MR analyses to assess for potential nonlinear J- or U-shape effects of LDL cholesterol lowering on AAA using the strategy of conditioning on quantiles of instrumental variable (IV)–free exposure and generating localized average causal effect (LACE) estimates (Burgess *et al.*, 2014; Sun *et al.*, 2019). 304,652 European-ancestry individuals in MVP were stratified into quintiles based on residual LDL-C after accounting for the effects of a PCSK9-LDL PRS (**Table S27**). Within each strata, ratio of effects MR was performed to test the effects of PCSK9-mediated increases in LDL-c on AAA risk. Two nonlinear P values based on the linear trend were estimated, and we set a $P < 0.05$ for statistical significance. Resultant odds ratios are scaled to relative PCSK9 inhibitor treatment effects based on clinical trial data.

### Banner authors

#### DiscovEHR

Regeneron personnel:
Goncalo Abecasis, Aris Baras, Michael Cantor, Giovanni Coppola, Aris Economides, Luca A. Lotta, John D. Overton, Jeffrey G. Reid, Alan Shuldiner, Andrew Deubler, Katia Karalis, Christina Beechert, Caitlin Forsythe, Erin D. Fuller, Zhenhua Gu, Michael Lattari, Alexander Lopez, Thomas D. Schleicher, Maria Sotiropoulos Padilla, Karina Toledo, Louis Widom, Sarah E. Wolf, Manasi Pradhan, Kia Manoochehri, Ricardo H. Ulloa, Xiaodong Bai, Suganthi Balasubramanian, Leland Barnard, Andrew Blumenfeld, Gisu Eom, Lukas Habegger, Alicia Hawes, Shareef Khalid, Evan K. Maxwell, William Salerno, Jeffrey C. Staples, Ashish Yadav, Dadong Li, Marcus B. Jones, Lyndon J. Mitnaul, Jason Mighty, Andrew Deubler, Katia Karalis, Katherine Siminovitch

Geisinger personnel:
Lance J. Adams, Jackie Blank, Dale Bodian, Derek Boris, Adam Buchanan, David J. Carey, Ryan D. Colonie, F. Daniel Davis, Dustin N. Hartzel, Melissa Kelly, H. Lester Kirchner, Joseph B. Leader, David H. Ledbetter, Ph.D., J. Neil Manus, Christa L. Martin, Raghu P. Metpally, Michelle Meyer, Tooraj Mirshahi, Matthew Oetjens, Thomas Nate Person, Christopher Still, Natasha Strande, Amy Sturm, Jen Wagner, Marc Williams

#### Regeneron Genetics Center

**RGC Management and Leadership Team**
Goncalo Abecasis, D.Phil. , Aris Baras, M.D. , Michael Cantor, M.D. , Giovanni Coppola, M.D. , Andrew Deubler , Aris Economides, Ph.D. , Katia Karalis, Ph.D. , Luca A. Lotta, M.D., Ph.D. , John D. Overton, Ph.D. , Jeffrey G. Reid, Ph.D. , Katherine Siminovitch, M.D. , Alan Shuldiner, M.D.

**Sequencing and Lab Operations**
Christina Beechert , Caitlin Forsythe, M.S. , Erin D. Fuller , Zhenhua Gu, M.S. , Michael Lattari , Alexander Lopez, M.S., John D. Overton, Ph.D. , Maria Sotiropoulos Padilla, M.S. , Manasi Pradhan, M.S. , Kia Manoochehri, B.S. , Thomas D. Schleicher, M.S. , Louis Widom , Sarah E. Wolf, M.S. , Ricardo H. Ulloa, B.S.

**Clinical Informatics**
Amelia Averitt, Ph.D. , Nilanjana Banerjee, Ph.D. , Michael Cantor, M.D. , Dadong Li, Ph.D. , Sameer Malhotra, M.D. , Deepika Sharma, MHI , Jeffrey Staples , Ph.D.

**Genome Informatics**
Xiaodong Bai, Ph.D. , Suganthi Balasubramanian, Ph.D. , Suying Bao, Ph.D. , Boris Boutkov, Ph.D. , Siying Chen, Ph.D. , Gisu Eom, B.S. , Lukas Habegger, Ph.D. , Alicia Hawes, B.S. ,

Shareef Khalid , Olga Krasheninina, M.S. , Rouel Lanche, B.S. , Adam J. Mansfield, B.A. , Evan K. Maxwell, Ph.D. , George Mitra, B.A. , Mona Nafde, M.S. , Sean O'Keeffe, Ph.D. , Max Orelus, B.B.A. , Razvan Panea, Ph.D. , Tommy Polanco, B.A. , Ayesha Rasool, M.S. , Jeffrey G. Reid, Ph.D. , William Salerno, Ph.D. , Jeffrey C. Staples, Ph.D. , Kathie Sun, Ph.D. , Jiwen Xin, Ph.D.

**Analytical Genomics and Data Science**
Goncalo Abecasis, D.Phil. , Joshua Backman, Ph.D. , Amy Damask, Ph.D. , Lee Dobbyn, Ph.D. , Manuel Allen Revez Ferreira, Ph.D. , Arkopravo Ghosh, M.S. , Christopher Gillies, Ph.D. , Lauren Gurski, B.S. , Eric Jorgenson, Ph.D. , Hyun Min Kang, Ph.D. , Michael Kessler, Ph.D. , Jack Kosmicki, Ph.D. , Alexander Li , Ph.D. , Nan Lin, Ph.D. , Daren Liu, M.S. , Adam Locke, Ph.D. , Jonathan Marchini, Ph.D. , Anthony Marcketta, M.S. , Joelle Mbatchou, Ph.D. , Arden Moscati, Ph.D. , Charles Paulding, Ph.D. , Carlo Sidore, Ph.D. , Eli Stahl, Ph.D. , Kyoko Watanabe, Ph.D. , Bin Ye, Ph.D. , Blair Zhang, Ph.D. , Andrey Ziyatdinov, Ph.D.

**Therapeutic Area Genetics**
Ariane Ayer, B.S. , Aysegul Guvenek, Ph.D. , George Hindy, Ph.D. , Giovanni Coppola, M.D. , Jan Freudenberg, M.D. , Jonas Bovijn M.D. , Julie Horowitz, Ph.D. , Katherine Siminovitch, M.D. , Jonas B. Nielsen, MD, PhD, Kavita Praveen, Ph.D. , Luca A. Lotta, M.D. , Manav Kapoor, Ph.D. , Mary Haas, Ph.D. , Moeen Riaz , Ph.D. , Niek Verweij, Ph.D. , Olukayode Sosina, Ph.D. , Parsa Akbari, Ph.D. , Priyanka Nakka, Ph.D. , Sahar Gelfman, Ph.D. , Sujit Gokhale, B.E. , Tanima De, Ph.D. , Veera Rajagopal, Ph.D. , Alan Shuldiner, M.D. , Bin Ye, Ph.D. , Gannie Tzoneva, Ph.D. , Juan Rodriguez-Flores, Ph.D.

**RGC Biology**
Shek Man Chim, Ph.D. , Valerio Donato, Ph.D. , Aris Economides, Ph.D. , Daniel Fernandez, M.S. , Giusy Della Gatta, Ph.D. , Alessandro Di Gioia, Ph.D. , Kristen Howell, M.S. , Katia Karalis, Ph.D. , Lori Khrimian, Ph.D. , Minhee Kim, Ph.D. , Hector Martinez , Lawrence Miloscio, B.S. , Sheilyn Nunez, B.S. , Elias Pavlopoulos, Ph.D. , Trikaldarshi Persaud, B.S.

**Research Program Management & Strategic Initiatives**
Esteban Chen, M.S. , Marcus B. Jones, Ph.D. , Michelle G. LeBlanc, Ph.D. , Jason Mighty, Ph.D. , Lyndon J. Mitnaul, Ph.D. , Nirupama Nishtala, Ph.D. , Nadia Rana, Ph.D.

#### UK Aneurysm Growth Study

Sara Baker, Jamie Barwell, Marcus Brooks, Neil Browning, Ian Chetter, Sohail Choksy, Alun Davies, Mark Dayer, Jonothan Earnshaw, Louis Fligelstone, Mark Gannon, Eric Grocott, Paul Hayes, Chris Imray, Nilesh Samani, Tim Lees, Gabor Libertiny, Charles McCollum, Colin Nice, Rajiv Pathak, Arun Pherwani, Lynda Pike, John Quarmby, Thomas Rix, Rob Sayers, Cliff Shearman, Vince Smyth, Mike Sweeting, Tim Sykes, William Tennant, John Thompson, Rao Vallabhaneni, Syed Yusuf, Frank Dudbridge

#### DBDS Genomic Consortium

Steffen Andersen[1], Karina Banasik[2], Søren Brunak[2], Kristoffer Burgdorf[3], Maria Didriksen[3], Khoa Manh Dinh[4], Christian Erikstrup[4], Daniel Gudbjartsson[5], Thomas Folkmann Hansen[6], Henrik Hjalgrim[7], Gregor Jemec[8], Poul Jennum[9], Pär Ingemar Johansson[3], Margit Anita Hørup Larsen[3], Susan Mikkelsen[4], Kasper Rene Nielsen[10], Mette Nyegaard[11], Sisse Rye Ostrowski[3], Ole Birger Pedersen[12], Kari Stefansson[5], Hreinn Stefánsson[5], Susanne Sækmose[12], Erik Sørensen[3], Unnur Þorsteinsdóttir[5], Mie Topholm Brun[13], Henrik Ullum[14], Thomas Werge[15]

[1] Department of Finance, Copenhagen Business School, Copenhagen, Denmark
[2] Novo Nordisk Foundation Center for Protein Research, Faculty of Health and Medical Sciences, University of Copenhagen, Copenhagen, Denmark
[3] Department of Clinical Immunology, Copenhagen University Hospital – Rigshospitalet, Copenhagen, Denmark
[4] Department of Clinical Immunology, Aarhus University Hospital, Aarhus
[5] deCODE Genetics, Reykjavik, Iceland
[6] Danish Headache Center, Department of Neurology, Copenhagen University Hospital, Rigshospitalet – Glostrup
[7] Department of Epidemiology Research, Statens Serum Institut, Centre for Cancer Research, Danish Cancer Society, Copenhagen, Denmark
[8] Department of Clinical Medicine, Sealand University hospital – Roskilde, Roskilde, Denmark
[9] Department of clinical neurophysiology, University of Copenhagen, Copenhagen, Denmark
[10] Department of Clinical Immunology, Aalborg University Hospital, Aalborg, Denmark
[11] Department of Biomedicine, Aarhus University, Aarhus, Denmark
[12] Department of Clinical Immunology, Zealand University Hospital – Køge, Køge, Denmark
[13] Department of Clinical Immunology, Odense University Hospital, Odense, Denmark
[14] Statens Serum Institute, Copenhagen, Denmark
[15] Institute of Biological Psychiatry Mental Health Centre, Sct. Hans, Copenhagen University Hospital – Roskilde, Roskilde, Denmark

#### VA Million Veteran Program

**MVP Executive Committee**

- Co-Chair: J. Michael Gaziano, M.D., M.P.H.
- Co-Chair: Rachel Ramoni, D.M.D., Sc.D.
- Jean Beckham, Ph.D.
- Jim Breeling, M.D. (ex-officio)
- Kyong-Mi Chang, M.D.
- Grant Huang, Ph.D. (ex-officio)
- Sumitra Muralidhar, Ph.D.
- Christopher J. O'Donnell, M.D., M.P.H.
- JP Casas Romero, M.D., Ph.D., Ex-Officio
- Philip S. Tsao, Ph.D.

**MVP Program Office**

- Sumitra Muralidhar, Ph.D.
- Jennifer Moser, Ph.D.

**MVP Recruitment/Enrollment**

- Recruitment/Enrollment Director/Deputy Director, Boston – Stacey B. Whitbourne, Ph.D.; Jessica V. Brewer, M.P.H.
- MVP Coordinating Centers
    - Clinical Epidemiology Research Center (CERC), West Haven – John Concato, M.D., M.P.H.
    - Cooperative Studies Program Clinical Research Pharmacy Coordinating Center, Albuquerque - Stuart Warren, J.D., Pharm D.; Dean P. Argyres, M.S.
    - Genomics Coordinating Center, Palo Alto – Philip S. Tsao, Ph.D.
    - Massachusetts Veterans Epidemiology Research Information Center (MAVERIC), Boston - J. Michael Gaziano, M.D., M.P.H.
    - MVP Information Center, Canandaigua – Brady Stephens, M.S.
- Core Biorepository, Boston – Mary T. Brophy M.D., M.P.H.; Donald E. Humphries, Ph.D.
- MVP Informatics, Boston – Nhan Do, M.D.; Shahpoor Shayan
- Data Operations/Analytics, Boston – Xuan-Mai T. Nguyen, Ph.D.

**MVP Science**

- Genomics - Christopher J. O'Donnell, M.D., M.P.H.; Saiju Pyarajan Ph.D.; Philip S. Tsao, Ph.D.
- Phenomics - Kelly Cho, M.P.H, Ph.D.
- Data and Computational Sciences – Saiju Pyarajan, Ph.D.
- Statistical Genetics – Elizabeth Hauser, Ph.D.; Yan Sun, Ph.D.; Hongyu Zhao, Ph.D.

**MVP Local Site Investigators**

- Atlanta VA Medical Center (Peter Wilson)
  1670 Clairmont Rd, Decatur, GA 30033
- Bay Pines VA Healthcare System (Rachel McArdle)
  10,000 Bay Pines Blvd Bay Pines FL 33744
- Birmingham VA Medical Center (Louis Dellitalia)
  700 S. 19th Street Birmingham AL 35233
- Cincinnati VA Medical Center (John Harley)
  3200 Vine Street, Cincinnati, OH 45220
- Clement J. Zablocki VA Medical Center (Jeffrey Whittle)
  5000 West National Avenue, Milwaukee, WI 53295
- Durham VA Medical Center (Jean Beckham)
  508 Fulton Street Durham, NC 27705
- Edith Nourse Rogers Memorial Veterans Hospital (John Wells)
  200 Springs Road, Bedford, MA 01730

- Edward Hines, Jr. VA Medical Center (Salvador Gutierrez)
  5000 South 5th Avenue, Hines, IL 60141
- Fayetteville VA Medical Center (Gretchen Gibson)
  1100 N College Ave, Fayetteville, AR 72703
- VA Health Care Upstate New York (Laurence Kaminsky)
  113 Holland Avenue Albany NY 12208
- New Mexico VA Health Care System (Gerardo Villareal)
  1501 San Pedro Drive, S.E.Albuquerque, NM 87108
- VA Boston Healthcare System (Scott Kinlay)
  150 S. Huntington Avenue, Boston, MA 02130
- VA Western New York Healthcare System (Junzhe Xu)
  3495 Bailey Avenue Buffalo, NY 14215-1199
- Ralph H. Johnson VA Medical Center (Mark Hamner)
  109 Bee Street, Mental Health Research, Charleston, SC 29401
- Wm. Jennings Bryan Dorn VA Medical Center (Kathlyn Sue Haddock)
  6439 Garners Ferry Road, Columbia, SC 29209
- VA North Texas Health Care System (Sujata Bhushan)
  4500 S. LANCASTER ROAD, DALLAS, TX 75216
- Hampton VA Medical Center (Pran Iruvanti)
  100 Emancipation Drive, Hampton, VA 23667
- Hunter Holmes McGuire VA Medical Center (Michael Godschalk)
  1201 Broad Rock Blvd., Richmond, VA 23249
- Iowa City VA Health Care System (Zuhair Ballas)
  601 Highway 6 West, Iowa City, IA 52246-2208
- Jack C. Montgomery VA Medical Center (Malcolm Buford)
  1011 Honor Heights Dr., Muskogee, OK 74401
- James A. Haley Veterans' Hospital (Stephen Mastorides)
  13000 Bruce B. Downs Blvd., Tampa, FL 33612
- Louisville VA Medical Center (Jon Klein)
  800 Zorn Avenue, Louisville, KY 40206
- Manchester VA Medical Center (Nora Ratcliffe)
  718 Smyth Road, Manchester, NH 03104
- Miami VA Health Care System (Hermes Florez)
  1201 NW 16th Street, 11 GRC, Miami FL 33125
- Michael E. DeBakey VA Medical Center (Alan Swann)
  2002 Holcombe Blvd. Houston TX 77030
- Minneapolis VA Health Care System (Maureen Murdoch)
  One Veterans Drive Minneapolis MN 55417
- N. FL/S. GA Veterans Health System (Peruvemba Sriram)
  1601 SW Archer Road, Gainesville, FL 32608
- Northport VA Medical Center (Shing Shing Yeh)
  79 Middleville Road, Northport, NY 11768
- Overton Brooks VA Medical Center (Ronald Washburn)

510 East Stoner Ave, Shreveport, LA 71101

- Philadelphia VA Medical Center (Darshana Jhala)
  3900 Woodland Avenue, Philadelphia, PA 19104
- Phoenix VA Health Care System (Samuel Aguayo)
  650 E. Indian School Road, Phoenix, AZ 85012
- Portland VA Medical Center (David Cohen)
  3710 SW U.S. Veterans Hospital Road, Portland, OR 97239
- Providence VA Medical Center (Satish Sharma)
  830 Chalkstone Avenue, Providence, RI 02908
- Richard Roudebush VA Medical Center (John Callaghan)
  1481 West 10th Street, Indianapolis, IN 46202
- Salem VA Medical Center (Kris Ann Oursler)
  1970 Roanoke Blvd.,Salem, VA 24153
- San Francisco VA Health Care System (Mary Whooley)
  4150 Clement Street, San Francisco, CA 94121
- South Texas Veterans Health Care System (Sunil Ahuja)
  7400 Merton Minter Boulevard, San Antonio, TX 78229
- Southeast Louisiana Veterans Health Care System (Amparo Gutierrez)
  2400 Canal Street, New Orleans, LA 70119
- Southern Arizona VA Health Care System (Ronald Schifman)
  3601 S 6th Ave, Tucson, AZ 85723
- Sioux Falls VA Health Care System (Jennifer Greco)
  2501 W 22nd St, Sioux Falls, SD 57105
- St. Louis VA Health Care System (Michael Rauchman)
  915 North Grand Blvd., St. Louis, MO 63106
- Syracuse VA Medical Center (Richard Servatius)
  800 Irving Avenue, Syracuse, NY 13210
- VA Eastern Kansas Health Care System (Mary Oehlert)
  4101 S 4th Street Trafficway, Leavenworth, KS 66048
- VA Greater Los Angeles Health Care System (Agnes Wallbom)
  11301 Wilshire Blvd Los Angeles, CA 90073
- VA Loma Linda Healthcare System (Ronald Fernando)
  11201 Benton Street, Loma Linda, CA 92357
- VA Long Beach Healthcare System (Timothy Morgan)
  5901 East 7th Street   Long Beach CA 90822
- VA Maine Healthcare System (Todd Stapley)
  1 VA Center, Augusta, ME 04330
- VA New York Harbor Healthcare System (Scott Sherman)
  423 East 23rd Street   New York, NY 10010
- VA Pacific Islands Health Care System (Gwenevere Anderson)
  459 Patterson Rd, Honolulu, HI 96819
- VA Palo Alto Health Care System (Philip Tsao)
  3801 Miranda Avenue   Palo Alto, CA 94304-1290

- VA Pittsburgh Health Care System (Elif Sonel)
  University Drive, Pittsburgh, PA 15240
- VA Puget Sound Health Care System (Edward Boyko)
  1660 S. Columbian Way Seattle, WA 98108-1597
- VA Salt Lake City Health Care System (Laurence Meyer)
  500 Foothill Drive Salt Lake City, UT 84148
- VA San Diego Healthcare System (Samir Gupta)
  3350 La Jolla Village Drive, San Diego, CA 92161
- VA Southern Nevada Healthcare System (Joseph Fayad)
  6900 North Pecos Road, North Las Vegas, NV 89086
- VA Tennessee Valley Healthcare System (Adriana Hung)
  1310 24th Ave. South Nashville, TN 37212
- Washington DC VA Medical Center (Jack Lichy)
  50 Irving St, Washington, D. C. 20422
- W.G. (Bill) Hefner VA Medical Center (Robin Hurley)
  1601 Brenner Ave, Salisbury, NC 28144
- White River Junction VA Medical Center (Brooks Robey)
  163 Veterans Drive, White River Junction, VT 05009
- William S. Middleton Memorial Veterans Hospital (Robert Striker)
  2500 Overlook Terrace, Madison, WI 53705
